# Empowering the crowd: Feasible strategies for epidemic management in high-density informal settlements. The case of COVID-19 in Northwest Syria

**DOI:** 10.1101/2020.08.26.20181990

**Authors:** Alberto Pascual-García, Jordan Klein, Jennifer Villers, Eduard Campillo-Funollet, Chamsy Sarkis

**Affiliations:** Institute of Integrative Biology. ETH-Zürich. Zürich, Switzerland; Office of Population Research. Princeton University. Princeton, NJ, USA; Princeton Environmental Institute. Princeton University. Princeton, NJ, USA; Genome Damage and Stability Centre. University of Sussex. Brighton, United Kingdom; Pax Syriana Foundation. Valetta, Malta

**Keywords:** COVID 19, Epidemiology, community-led interventions, SIR models, Internally Displaced Persons, Syria, Refugees

## Abstract

More than 1 billion people live in informal settlements worldwide, where precarious living conditions pose unique challenges to managing a COVID-19 outbreak. Taking Northwest Syria as a case-study, we simulated an outbreak in high-density informal Internally Displaced Persons (IDP) camps using a stochastic Susceptible-Exposed-Infectious-Recovered model. Expanding on previous studies, taking social conditions and population health/structure into account, we modeled several interventions feasible in these settings: moderate self-distancing, self-isolation of symptomatic cases, and protection of the most vulnerable in “safety zones”. We considered complementary measures to these interventions that can be implemented autonomously by these communities, such as buffer zones, health-checks, and carers for isolated individuals, quantifying their impact on the micro-dynamics of disease transmission. All interventions significantly reduce outbreak probability and some of them reduce mortality when an outbreak does occur. Self-distancing reduces mortality by up to 35% if contacts are reduced by 50%. A reduction in mortality by up to 18% can be achieved by providing 1 self-isolation tent per 8 people. Protecting the most vulnerable in a safety zone reduces the outbreak probability in the vulnerable population and has synergistic effects with the other interventions. Our model predicts that a combination of all simulated interventions may reduce mortality by more than 90% and delay an outbreak’s peak by almost two months. Our results highlight the potential for non-medical interventions to mitigate the effects of the pandemic. Similar measures may be applicable to controlling COVID-19 in other informal settlements, particularly IDP camps in conflict regions, around the world.

**Key questions:** *What is already known?:*

- Since the onset of the COVID-19 pandemic, many studies have provided evidence for the effectiveness of strategies such as social distancing, testing, contact tracing, case isolation, use of personal protective equipment/facemasks and improved hygiene to reduce the spread of the disease. These studies underlie the recommendations of the World Health Organisation, but their implementation is contingent on local conditions and resources.
- Mathematical modelling is the basis of many epidemiological studies and has helped inform policymakers considering COVID-19 responses around the world. Nevertheless, only a limited number of studies have applied these models to informal settlements.

*What are the new findings?:*

- We developed a mathematical model to study the dynamics of COVID-19 in Syrian IDP camps, elaborating on previous efforts done in similar settings by explicitly parameterizing the camps’ demographics, living conditions and micro-dynamics of interpersonal contacts in our modelization.
- We designed interventions such as self-distancing, self-isolation and the creation of safety zones to protect the most vulnerable members of the population, among others, through conversations with camp managers with on-the-ground knowledge of what interventions would be feasible and have community buy-in.
- Our results show how low-cost, feasible, community-led non-medical interventions can significantly mitigate the impact of COVID-19 in Northwest Syrian IDP camps.

*What do the new findings imply?:*

- Our model represents a step forward in the much-needed search for epidemiological models that are sufficiently flexible to consider specific social questions. The model can also help inform similar interventions in refugee camps in conflict-torn regions, and potentially be adapted to other informal settlements and vulnerable communities around the world.

## Introduction

The spread of airborne infectious diseases with pandemic potential in regions immersed in protracted armed conflicts, where large portions of their populations have become displaced, is an important challenge [1]. When the displaced population exceeds official resettlement and refugee camp capacity, Internally Displaced Persons (IDPs) must live in informal settlements (hereafter named “camps”). These regions must contend with the public health challenges resulting from violence [2], the deterioration of health-systems [3], especially of critical care [4], and the breakdown of essential public infrastructure such as water and sanitation systems [5]. Urgent action is needed to contain the spread of disease in these settings, a task which necessarily involves the engagement of the communities living in them [6].

This study focuses on the spread of COVID-19 in Northwest region of Syria (NWS): a relatively small geographical area with 4.2 million people, of which 1.15 million (27.4%) are IDPs living in camps [7], and where the number of cases increased twenty-fold between September 8th and October 20th, 2020 [8]. The health status of households in camps in NWS is poor; 24% have a member with a chronic disease, of whom 41% have no access to medicines [9]. As in other conflict regions, the political instability in NWS hinders coordinated public health actions, and the ongoing movements of IDPs create ample opportunity for infectious disease transmission, while making contact tracing interventions infeasible.

To investigate feasible COVID-19 prevention interventions in the camps, we considered a Susceptible-Exposed-Infectious-Recovered model similar to the one presented by Gatto *et al*. [10], in which the camps’ populations are divided into classes reflecting their estimated age-structures and comorbidity prevalence. We use this model to propose various interventions aimed at reducing the number of contacts within and between population classes in general, and with symptomatic individuals in particular. We paid special attention to how the living conditions in informal camps inform the assumptions underlying our proposed interventions, a question often neglected [11]. We modeled interventions previously proposed for African cities [12], such as self-distancing, isolation of symptomatic individuals and the creation of a “safety zone” in which more vulnerable members of the population are protected from exposure to the virus.

Building upon the approach used to model the impact of these interventions in African cities, our model includes a parameterization of the contacts each individual has per day [12]. We further elaborate upon this approach by making a more explicit representation of contacts and other parameters in the model. We consider the micro-dynamics of contacts, the time that individuals take to recognize their symptoms before self-isolating, the effect of having carers to attend to isolated individuals, and the existence of a buffer zone in which exposed and protected population classes can interact under certain rules. We examine a potential worst-case scenario in which there is no access to any healthcare facility. Since empowering local communities in conflict regions to understand how to control diseases like COVID-19 is possibly the most (and perhaps only) effective way to minimize its spread, our models are of utmost importance for informing the implementation of realistic interventions in these regions.

## Methods

### The model

We consider a model simulating a viral outbreak in a single camp over a 12-month period inspired by those proposed by Gatto *et al*. and Bertuzzo *et al*. [10, 13] (see Fig. 1). The model is adapted to the context of NWS IDP camps and is divided into compartments containing individuals at different possible stages along the disease’s progression, governed by the following set of differential equations:

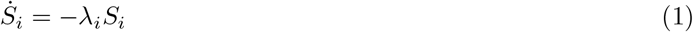

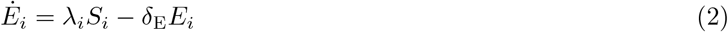

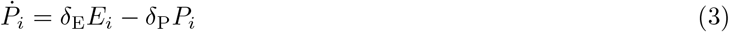

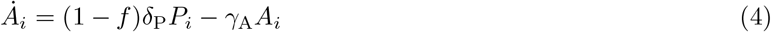

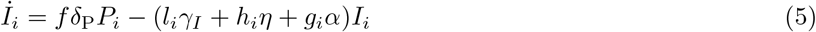

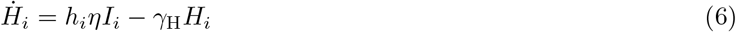

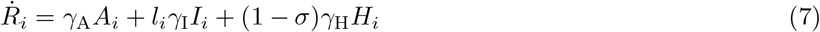

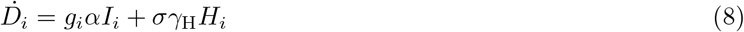

**Figure 1:**
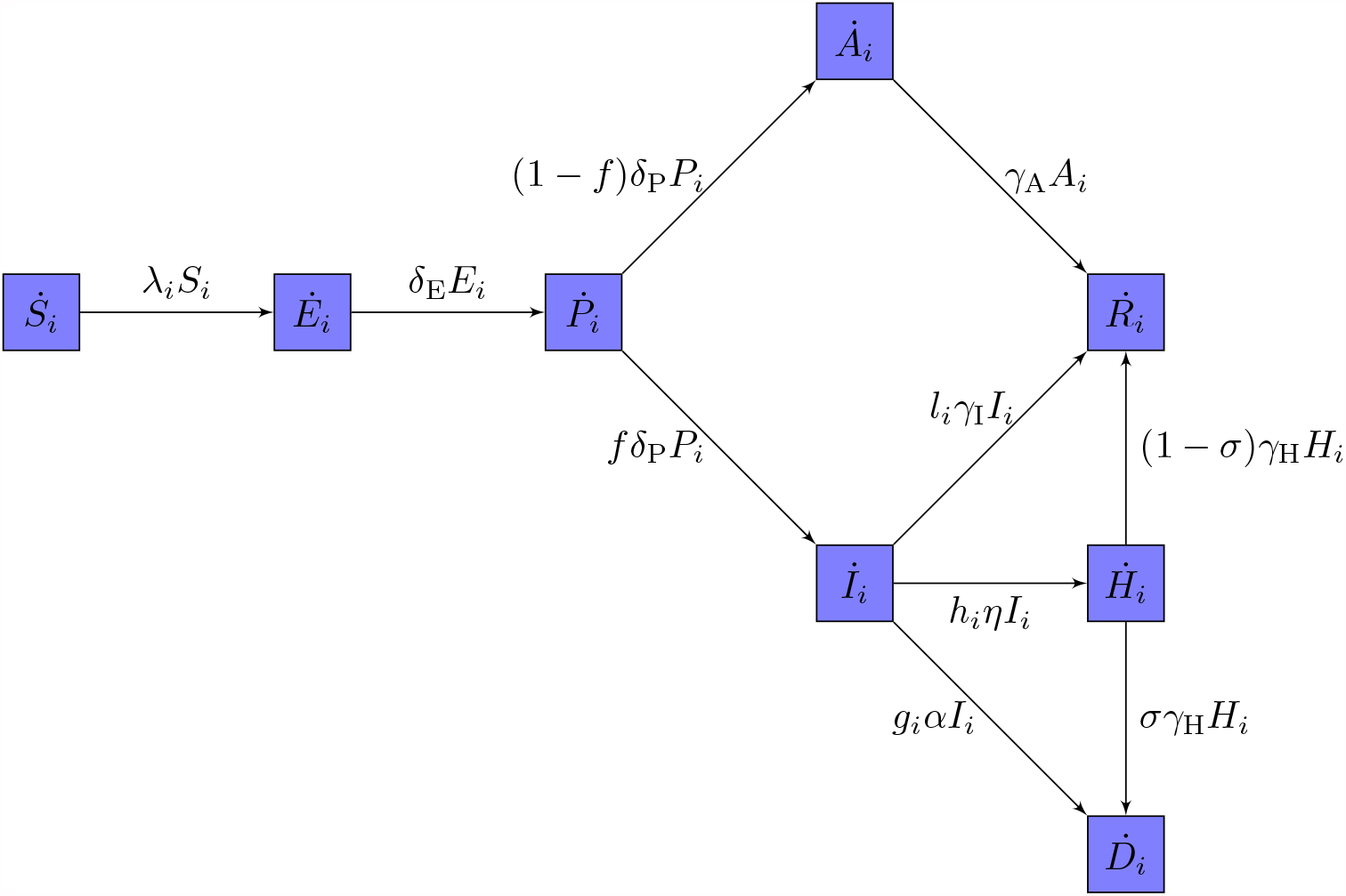
Diagram of the model. The model considers the following compartments: susceptible (S), exposed (E), infectious-presymptomatic (P), infectious-asymptomatic (A), infectious-symptomatic (I), infectious-requiring hospitalization (H), recovered (R) and dead (D).

The susceptible population (*S*_*i*_) becomes exposed at rate *λ*_*i*_, while exposed individuals (*E*_*i*_) progress through the latent period at rate *δ*_E_ to a preclinical infectious stage (*P*_*i*_), which then progresses to (at rate *δ*_P_) either a a clinical (symptomatic, *I*_*i*_, with probablity *f*) or subclinical (asymptomatic, *A*_*i*_, with probability 1 – *f*) infectious stage. Asymptomatic cases recover (*R*_*i*_) at rate *γ*_A_. Symptomatic cases have 3 potential outcomes: mild cases will recover at rate *γ*_I_, severe cases will progress to an extended infectious period during which they require hospitalization (*H*_*i*_) at rate *η*, while critical cases requiring intensive care unit (ICU) care will die (*D*_*i*_) at rate *α*. Finally, since the fate of individuals in the hospitalized compartment is uncertain if healthcare is not available, we run simulations considering two possibilities: either all recover (*σ* = 0), or all die (*σ* = 1) (see section Epidemiological severity assumptions). The specific values for the parameters are presented in Table 1.

**Table 1:** Fixed parameters. See Supplementary Materials for details.

| Parameter | Description | Value | Distribution | Reference |
| --- | --- | --- | --- | --- |
| $1/\delta_E + 1/\delta_P$ | Incubation period (days) | 5.2 (95% CI: 4.1-7.0) | Lognormal | [17] |
| $1/\delta_P$ | Presymptomatic infectious period (days) | 2.3 (95% CI: 0.8-3.8) | Gaussian | [18, 19] |
| $1/\delta_E$ | Latent period (days) | $(1/\delta_E + 1/\delta_P) - 1/\delta_P$<br>(Minimum = .5 days) | | Derived |
| $1/\gamma_I$ | Symptomatic infectious period (days) | 7 | --- | [18, 20] |
| $1/\gamma_A$ | Asymptomatic infectious period (days) | 7 | --- | [18, 20] |
| $1/\eta$ | Time from symptom onset to requiring hospitalization (days) | 7 (IQR: 4-8) | Gamma | [21] |
| $1/\alpha$ | Time from symptom onset to death (critical cases, days) | 10 (IQR: 6-12) | Gamma | [21] |
| $1/\gamma_H$ | Time from requiring hospitalization to recovery/death (days) | 10 (IQR: 7-14) | Gamma | [21] |
| $f$ | Probability an infectious individual is symptomatic | 0.84 (95% CI: 0.8-0.88) | Binomial | [22] |
| $\sigma$ | Indicator of whether hospitalized recover or die | $\sigma \in \{0, 1\}$ | --- | Assumed |

While we introduced the model as a classical system of ordinary differential equations (Eqs. 1–8) we considered a stochastic implementation [14], with an integer description of the population in which a state is encoded in a vector *X* = (*S, E, P, I, A, H, R, D*)^*t*^, with the total population size conserved throughout the simulation *N* = *S* + *E* + *P* + *I* + *A* + *H* + *R* + *D*. The possible transitions between states of the system are shown in Fig. 1, where an arrow indicates transitions in which the source compartment transfers one individual to the target compartment. The mean transition rates corresponding to each transition are displayed. The system then evolves following a continuous-time Markov process which is simulated following the Gillespie algorithm implemented in the adaptivetau package [15].

### Demographic– and behaviour-classes

The model splits the population into classes (indexed *i*) to account for heterogeneity with respect to clinical risk and behaviour. Working with population classes allows us to encode behavioural assumptions in the model and strike an appropriate balance between generality, computational tractability, and the requisite specificity to realistically evaluate our proposed interventions [16]. Moreover, the explicit representation of contacts between and within population classes allows us to design interventions considering cultural and context-specific assumptions [11] (see section Interventions and Supplementary Material for details). Under a null model where no interventions are implemented, the distinctions between classes are only dependent on age and comorbidity status (hereafter “demographic-classes”). *h*_*i*_, *g*_*i*_ and *l*_*i*_ are demographic-class specific parameters, adjusted to ensure that the proportions of symptomatic cases progressing through each of the 3 potential clinical outcomes (mild, severe, and critical) are consistent with the literature (see section Epidemiological severity assumptions).

Under some interventions, the demographic-classes may be subdivided further into subclasses according to behaviour (“behaviour-classes”). Consequently, different interventions may require models with different numbers of classes. We refer to both demographic- and behaviour-classes generically as “classes” (see section Interventions for the modelization of behaviour-classes).

### Population structure of demographic-classes

We parameterized the model with data from IDPs in NWS [23]. The population sizes of informal camps are right-skewed, with a mean of 1212. We simulated camps with populations of 500, 1000 and 2000 individuals. Since interventions tend to be less effective in larger camps, the results presented refer to simulations with 2000 individuals, unless otherwise specified. For our demographic-classes, we considered 3 age groups: children (age 1, 0-12 years old), younger adults (age 2, 13-50 yrs) and older adults (age 3, *>*50 yrs). For ages 2 and 3, we considered two subclasses comprising healthy individuals and individuals with comorbidities (see Table 2).

**Table 2:** Demographic class-specific parameters. Estimated proportions of individuals in the population and mean number of contacts per individual per day for each demographic class. See Supplementary Materials for derivations.

| Parameter | Description | Demographic-class |  |  |  |  | References |
| --- | --- | --- | --- | --- | --- | --- | --- |
|  |  | Age 1<br>(0-12) | Age 2 (13-50)<br>no comorbidities | Age 2 (13-50)<br>comorbidities | Age 3 (>50)<br>no comorbidities | Age 3 (>50)<br>comorbidities |  |
| Fraction<br>in class | - | 0.407 | 0.471 | 0.0626 | 0.022 | 0.0373 | [23, 24] |
| $c_i$ | Mean<br>contacts<br>per day | 25 | 15 | 15 | 10 | 10 | From<br>camp<br>managers |

### Transmissibility assumptions

Although individuals in IDP camps share tents with other co-occupants, whom they may be more likely to infect than occupants of different tents, we ignore spatial structure in our model and assume a well-mixed population.

This is justified because individuals from different tents share common spaces (e.g. latrines) and have frequent interactions with each other, especially among children. Consequently, our following derivation of the transmissivity parameter itself, *τ*, is not spatially explicit.

The rate at which susceptible individuals become exposed is

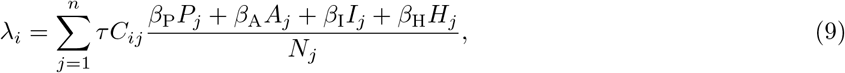

where *C*_*ij*_ is the average number of contacts that individuals of class *i* have with individuals of class *j* per day and *N*_*j*_ is the total population size of class *j*. We parameterized *C*_*ij*_ by multiplying the mean number of total contacts that individuals from a population class *i* have per day, *c*_*i*_, by the probability of random interaction with individuals of class *j*. Considering a well-mixed population, this probability is proportional to class *j*’s fraction of the total population, i.e. *C*_*ij*_ = *c*_*i*_*N*_*j*_*/N*. If interventions are absent, we consider demographic-classes only and, hence, different values of *c*_*i*_ reflect heterogeneity in the number of contacts by demographic-class. We assume specific values of *c*_*i*_ for each class based on conversations with camp managers in NWS (see Table 2).

The probability of infection if there is a contact between a susceptible and an infected person is *τβ*_P_, *τβ*_A_, *τβ*_I_ or *τβ*_H_ depending upon whether the infected individual is in the presymptomatic (*P*_*i*_), symptomatic (*I*_*i*_), asymp-tomatic (*A*_*i*_), or hospitalized compartment (*H*_*i*_), respectively. The *τ* parameter is the maximum transmissivity, which is observed at the presymptomatic stage for individuals who go on to become symptomatic [18]. Thus, we selected the transmissivity of these individuals as a reference (*β*_P→I_ = 1) with the remaining *β* parameters set relative to *β*_P→I_ (*β*_*i*_ *< β*_P→I_, *i ϵ* {P, A, H, I}), where the mean transmissibility of all presymptomatic individuals (*β*_P_) is estimated as a weighted average of the transmissibility of individuals that will become symptomatic and asymptomatic (see Table 3, Supplementary Materials for derivation).

**Table 3:** Transmissibility parameters. See Supplementary Materials for details.

| Parameter | Description | Value | Distribution | Reference |
| --- | --- | --- | --- | --- |
| $\tau$ | Maximum transmissibility | 0.14 (95% CI: 0.05-0.40) | Lognormal | Derived |
| $\beta_{P \rightarrow I}$ | Presymptomatic transmissibility of individuals becoming symptomatic relative to $\tau$ | Reference stage (=1) | --- | [28, 19] |
| $\beta_P$ | Mean presymptomatic transmissibility relative to $\tau$ | 0.93 (95% CI: 0.88-0.99) | Empirical | Derived |
| $\beta_A$ | Asymptomatic transmissibility relative to $\tau$ | 0.14 (95% CI: 0.05-0.40) | Lognormal | Derived |
| $\beta_I$ | Clinical symptomatic transmissibility relative to $\tau$ | 0.24 (95% CI: 0.11-0.60) | Lognormal | Derived |
| $\beta_H$ | Hospitalized transmissibility relative to $\tau$ | 0.11 (95% CI: 0.05-0.29) | Lognormal | Derived |

The *τ* parameter was estimated by randomly generating a value for the basic reproduction number, *R*_0_, following a Gaussian distribution with a mean of 4 (99% CI: 3–5) and dividing this value by the the dominant eigenvalue of the Next Generation Matrix (see section Computational implementation for details and Supplementary Material for the analytical results). The distribution of *R*_0_ was a compromise between values reported in the literature from regions with high-density informal settlements: *R*_0_=2.77 in Abuja and 3.44 in Lagos, Nigeria [25], 3.3 in Buenos Aires [26], and 5 in Rohingya refugee camps in Bangladesh [27].

### Epidemiological severity assumptions

In NWS, there are 4 active and 2 planed COVID-19 referral hospitals, with a current capacity of 66 ventilators, 74 ICU beds and 355 ward beds for 4.2 million people [29, 30]. Estimations based on an exponential growth model from Hariri et al. predicted a collapse of health facilities 8 weeks into an outbreak [31]. Although we do not have access to official data on healthcare occupancy, the currently reported number of cases suggests that this scenario could have been reached [8]. Hence, we considered a worst-case scenario in which individuals will not have access to healthcare and assumed that all critical cases (those requiring ICU care) would die. However, there is greater uncertainty about the fate of severe cases, those requiring hospitalization but not ICU care. We therefore considered a compartment for severe cases to account for a longer infectious period if they stay in the camp (see compartment *H*_*i*_, Fig. 1). This compartment also helped us model some interventions more realistically, for example by noting that the symptoms of severe cases are incompatible with self-isolation. To estimate upper and lower bounds for the outcome variables of our model, we simulated two possible scenarios for the fate of this compartment: one in which all cases recover, and another in which all die. In the simulations presented in the Main Text, we consider the worst-case scenario in which all of these cases die.

The fractions of symptomatic cases that are severe 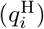, critical 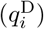 and recover (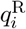, where 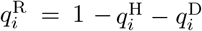) are demographic-class-specific (see Table 4). We estimated the fractions of symptomatic cases in each demographic-class that would become severe 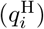 and critical (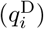) using data from developed countries with superior population health [32, 33]. Following previous work [12], we mapped the age-specific case severity distributions of the NW Syrian adult population to those of 10 years older age groups in developed countries.

**Table 4:** Proportion of symptomatic cases that become severe and critical. Since the rates at which these cases become severe, critical, or recover are different, we introduced three paramaters (*h*_*i*_, *g*_*i*_ and *l*_*i*_) to distribute individuals according to the desired proportions (values between parenthesis). The proportion of individuals recovering is computed as 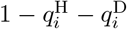 (see Supplementary Materials for details).

| Parameter | Description | Demographic class |  |  |  |  |
| --- | --- | --- | --- | --- | --- | --- |
|  |  | Age 1 (0-12) | Age 2 (13-50)<br>no<br>comorbidities | Age 2 (13-50)<br>comorbidities | Age 3 (>50)<br>no<br>comorbidities | Age 3 (>50),<br>comorbidities |
| $q_i^H (h_i)$ | Fraction of symptomatic cases severe | 0.064 (0.064) | 0.067 (0.066) | 0.199 (0.191) | 0.183 (0.178) | 0.445 (0.406) |
| $q_i^D (g_i)$ | Fraction of symptomatic cases critical | 0.0065 (0.009) | 0.020 (0.028) | 0.094 (0.129) | 0.063 (0.088) | 0.222 (0.289) |

Since the rates at which clinical symptomatic individuals (*I*_*i*_) resolve into these three epidemiological outcomes are different (*η* for *H, α* for *D* and *γ*_I_ for *R*) we introduced three parameters, *h*_*i*_, *g*_*i*_ and *l*_*i*_, to distribute individuals according to the desired proportions. The analytic solution is provided in Suppl. Material and the specific values in Table 4.

### Interventions

The interventions we consider are modelled by modifying the rate at which individuals become exposed (the term *λ*_*i*_, Eq. 9), and/or adding new population classes that govern behavioural changes (behaviour-classes). Since *λ*_*i*_ can be factorized in four terms, the interventions may influence one or several of these terms. The factors present in *λ*_*i*_ and the terms modulating them in the interventions are (see Eq. 10): i) the maximum transmissibility, *τ*, which is reduced in some interventions by a factor *ξ*_*ij*_, when interactions are restricted to buffer zones (see below for details); ii) the average number of contacts that individuals in class *i* have per day, *c*_*i*_. This quantity can either be uniformly reduced across all classes, or the contact rate of class *i* with class *j* can be modified. We model this modification of contact rates using the matrix *ϵ*_*ij*_; iii) the probability of encounter between members of class *i* and *j* in a well-mixed population, *N*_*j*_*/N*. This probability can vary by modifying the visibility of a member of class *j* to a member of class *i*, which we express with the matrix *ω*_*ij*_; and iv) the probablility of becoming infected by individuals at specific stages of the disease (e.g. for hospitalized individuals this is encoded in the term *β*_H_*H*_*j*_*/N*_*j*_) can also be modified by specific factors. Only the terms for individuals at the clinical symptomatic (*I*) and hospitalized (*H*) stages are modified in our interventions, through the parameters *ζ*_I_ and *ζ*_H_, respectively. Following these considerations, the generic form of *λ*_*i*_ under the interventions becomes:

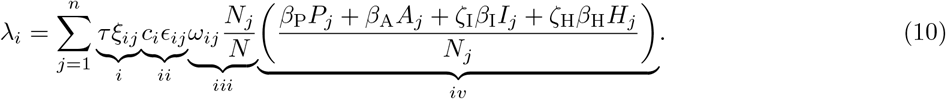

All interventions, self-distancing (see Fig. 2-1), self-isolation (see Fig. 2-2), safety zone (see Fig. 2-3), and evacuation (see Fig. 2-4) can be parameterized following this expression. The specific values of the parameters are presented in Table 5 and their derivations in Supplementary Materials.

**Table 5:** Parameterization of the interventions. Values of *ϵ*_*ij*_, *ω*_*ij*_, *ζ*_I_, *ζ*_H_ and *ξ*_*ij*_ considered in the interventions (see Supplementary Materials for their derivations). *c*_care_ = number of contacts per day of each isolated individual with carers. *Ĩ*_*j*_ = Number of isolated cases of class *j. N*_I_ = number of symptomatic individuals. *Ñ* = maximum number of tents available for self-isolation. Θ(*x*) = Heaviside function. *c*_visit_ = maximum number of contacts that individuals in the green zone can have with individuals from the orange zone per day 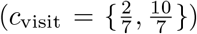. Under a “lockdown” of the green zone where visits are reduced by 50% 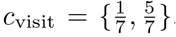, and where visits are reduced by 90% 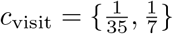. *ϑ* = fraction of the population in the orange (green) zone capable of coming in contact with individuals from the green (orange) zone. *ϑ* = 1 if class *i* is in the green zone, and is proportional to *c*_visit_*N*_g_*/N*_o_ if in the orange zone. *N*_X_ and *N*_Y_ generically refer to the total population in either the green (*N*_g_) or the orange (*N*_o_) zones. Note (*): The parameters *ζ*_I_ and *ζ*_H_ are set to 0 if health checks to access to the buffer zone are implemented in the intervention, and set to 1 otherwise.

| Intervention | $\epsilon_{ij}$ | $\omega_{ij}$ | $\zeta_I$ | $\zeta_H$ | $\xi_{ij}$ | Details |
| --- | --- | --- | --- | --- | --- | --- |
| Self-distancing | $\epsilon \in \{0.9, 0.8, 0.7, 0.6, 0.5\}$ | 1 | 1 | 1 | 1 | constant $\forall i, j$ |
| Self-isolation | $(c_{\text{care}}/c_i)(\tilde{I}_j/N_i)$ | $N/N_j$ | 1 | 1 | 0.2 | $i \in$ young healthy adults<br>$j \in$ isolated individuals in class $j$ |
| | 1 | 1 | $\Theta(N_I - \tilde{N})$ | | 1 | $i \in$ young healthy adults<br>$j \in$ individuals not isolated in class $j$ |
| | 0 | 0 | | | 0 | $i \notin$ young healthy adults<br>$j \in$ isolated individuals in class $j$ |
| | 1 | 1 | | | 1 | $i \notin$ young healthy adults<br>$j \in$ indiv in class $j$ |
| Safety zone | $\vartheta c_{\text{visit}}/c_i$ | $N/N_Y$ | $\{0,1\}$<br>see (*) | $\{0,1\}$<br>see (*) | 0.2 | $i \in$ zone $X, j \in$ zone $Y$<br>($X, Y \in \{\text{green, orange}\}$) |
| | $1 - \vartheta c_{\text{visit}}/c_i$ | $N/N_X$ | 1 | 1 | 1 | $i, j \in$ zone $X$<br>($X \in \{\text{green, orange}\}$) |
| Evacuation | 1 | 1 | 1 | 0 | 1 | — |

**Figure 2:**
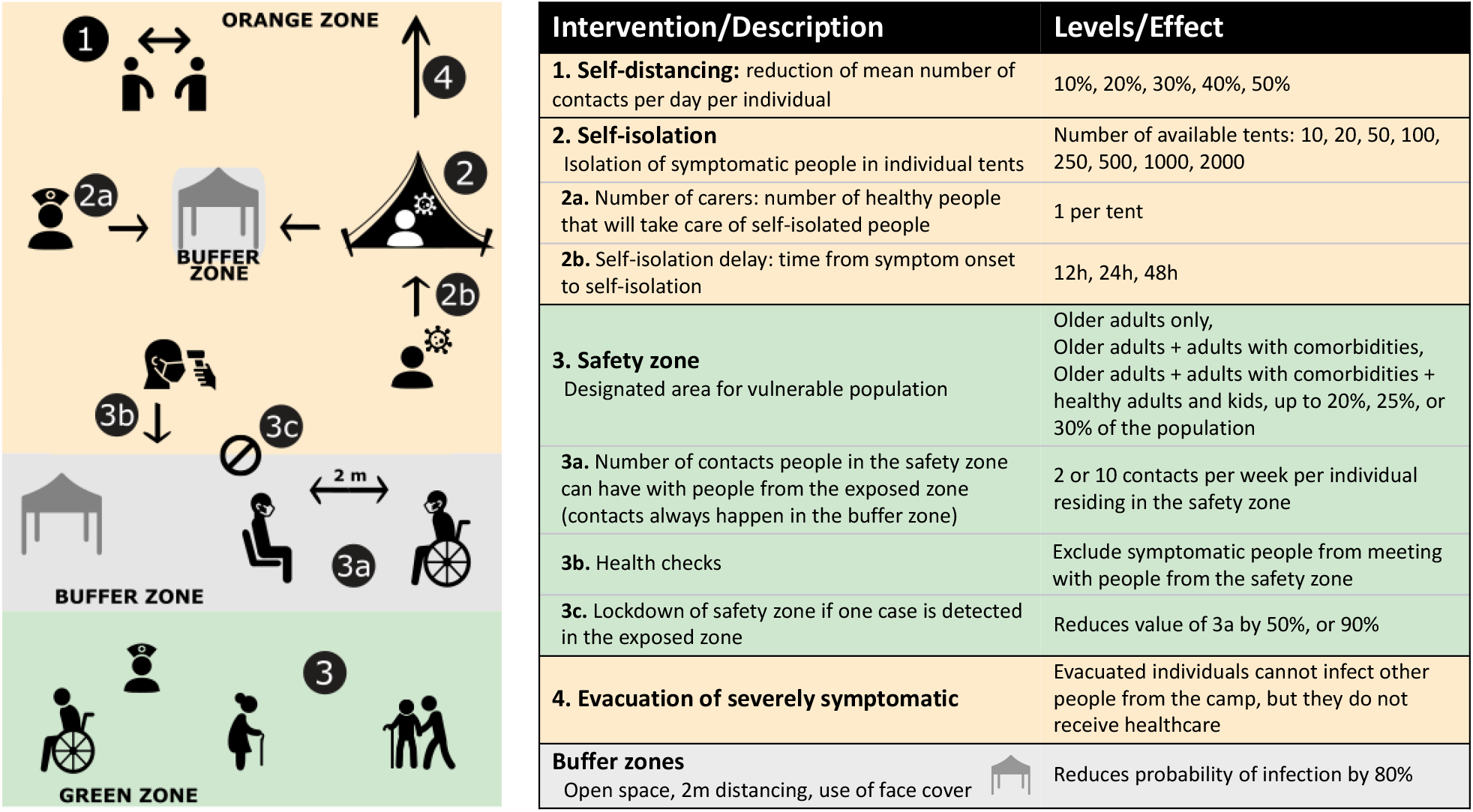
Diagram of interventions.

#### Self-distancing

The first non-medical intervention that we modeled is a reduction in the mean number of contacts per individual per day for the whole camp population (see Fig. 2-1). The average number of contacts of each class *c*_*i*_ (see Table 2) is reduced by a class-independent factor, hence the matrix *ϵ*_*ij*_ is uniform for all classes, i.e. *ϵ*_*ij*_ = *ϵ, ϵ* ∈ {0.9, 0.8, 0.7, 0.6, 0.5}. No further adjustments are required for this intervention. Since the mean number of inhabitants per tent in a camp is 5.5 and sanitation facilities are shared [23], we inferred that the number of contacts per day cannot be reduced by more than 50% (*ϵ* = 0.5). For a younger adult, this would mean 7.5 contacts per day.

#### Self-isolation

Self-isolation is a challenge in informal settlements, where households consist of a single (often small) space, water is collected at designated locations, sanitation facilities are communal and food supplies are scarce. We considered the possibility of those showing symptoms (i.e. in compartment I) self-isolating in individual tents in dedicated parts of the camps. We excluded individuals with severe symptoms (H compartment) from the intervention since they require additional care not compatible with self-isolation. Instead, we considered the possibility of evacuating these individuals from the camp as an additional intervention (see below). We simulated self-isolation with various numbers of isolation tents per camp, ranging from 10 to 2000 for a camp of 2000 people (see Fig. 2-2). In addition, we modeled the role of carers dedicated to providing for isolated individuals (see Fig. 2-2a). Carers are drawn from the younger adults class with no comorbidities who are not at a stage of the disease during which they display symptoms (i.e. not in compartments I or H), while isolated individuals may belong to any class. Since this intervention only modifies the infectiousness of individuals in the I compartment and the exposure of carers, the consideration of additional behavioural-classes is not required. Instead, the intervention can be encoded in *λ*_*i*_ by simply splitting the contribution of symptomatic individuals to the rate of exposure into two terms, one for the isolated individuals and another for the remaining population. Hence, its implementation requires deriving the parameters *ϵ*_*ij*_ and *ω*_*ij*_ for interaction between healthy younger adults (from which carers are drawn), the remaining classes, and isolated and non-isolated individuals separately. In Table 5 we present these terms and their derivation in Supplementary Materials.

In addition, interactions between carers and isolated individuals were restricted to *buffer zones*, which we envisioned as open spaces, with guidelines in place to limit occupancy to 4 individuals wearing masks with at least 2 meters of distance between them, where we assume transmissivity is reduced by 80% (ξ_*ij*_ = 0.2). In considering one carer per isolated individual with one contact per day, we do not neglect their probability of infecting the rest of the camp.

#### Safety zone

In this intervention, the camp is divided in two areas: a safety zone, in which more vulnerable people live (hereby referred to as a “green” zone following previous studies [12]), and an exposed (“orange”) zone with the remaining population. In our simulations, the first exposed individual always belongs to the orange zone. The living conditions within both zones remain the same, so the overall contact rate does not change unless self-distancing is also implemented. A consequence of maintaining the overall contact rate is that, reducing contacts with individuals living in a different zone, implies an increase in contacts with individuals in the same zone (see Supplementary Material). Although we do not expect this assumption to be true in general, it allows us to investigate undesired side-effects of this intervention, such as older adults having increased contacts amongst themselves if isolated together. Since proposals for partitioning the population may be received differently across camps, we considered several scenarios for allocating a camp population to the two zones (see Fig. 2-3). Implementing this intervention thus requires the split of some demographic-classes into two behaviour-classes, depending upon the scenario. For example, if some healthy younger adults are allocated into the green zone, we split the demographic-class “healthy younger adults” into “orange” and “green” behaviour-classes, to model the different contact rates that these two subclasses of healthy younger adults will have amongst themselves and with other classes. In Supplementary Table 2 we present the classes considered in each scenario.

Interactions between the two zones are limited to a buffer zone, reducing transmissivity (i.e. ξ_*ij*_ = 0.2, see previous section). Individuals in the green zone cannot leave and thus need to be provided with supplies by individuals in the orange zone, which will take place in the buffer zone. In our simulations, we considered limiting individuals in the green zone to 10 or 2 contacts with individuals from the orange zone per week (see Fig. 2-3a). Other variations of this intervention we explored include preventing symptomatic individuals from entering the buffer zone (incorporating health-checks, see Fig. 2-3b) and a “lockdown” of the green zone, where the number of weekly contacts in the buffer zone is reduced by 50% or 90% (see Fig. 2-3c). Although overall contact rates are conserved in this intervention, we modify the contact rates between *i* and *j* with *ϵ*_*ij*_ and the probability of interaction between *i* and *j* with ω_*ij*_, where both parameters are determined by whether *i* and *j* are in the same or different zones (see Table 5 for specific values and Supplementary Materials for derivation).

#### Evacuation

The last intervention we simulated is the evacuation of severe cases (individuals in the hospitalization compartment). Since they require more intensive care that cannot be delivered while adhering to the guidelines of a buffer zone, severe cases were assumed to be fully infectious and not able to self-isolate. Once severe cases are evacuated, their infectivity is reduced to zero (*ζ*_H_ = 0, see Fig. 2-4). The fate of severe cases is not altered by this intervention since we assumed that hospitals are saturated and that evacuees are transferred to isolation centers instead.

### Computational implementation and statistical analysis

The specific values of the parameters shown in Eqs. 1–8 and of *R*_0_ are independently drawn at each integration step from the probability distributions shown in Tables 1, 3 and 4. We note that by generating transition rates from the empirically-determined residence times, we are not reproducing these residence times for the simulated individuals. In our simulations, individuals will experience exponentially-distributed residence times with a mean equal to the mean of the correspondent distribution (to 1*/κ* in the case of the symptomatic compartment). The generation of random values around the mean following the empirical distribution at each time-step is aimed at adding noise to the mean, to partially account for the empirical uncertainty.

The Next Generation Matrix is also computed at each integration step from the parameters drawn, and *τ* estimated. In the code provided it is possible to fix the seed to exactly reproduce the results presented. Our simulations start with a completely susceptible population where one person in the younger adult population is exposed to the virus (who is also in the orange zone if the safety zone intervention is in place). We verified that a steady state was always reached before the end of each simulation. We did not consider migration, births, nor deaths due to other causes, since they are small enough in magnitude to not significantly impact the course of an outbreak, provided additional conflict does not erupt.

For each implementation of the interventions, we ran 2500 simulations and compared results between them. The main variables considered are the fraction of simulations in which at least one death is observed, a proxy for the probability of an outbreak, the fraction of the population that dies and the time until the symptomatic population peaks, as well as the infection fatality rate (IFR), the fractions of the population that are recovered and remain susceptible at steady-state. For consistency, we only considered simulations in which there was an outbreak when comparing the outcome of a variable between interventions. We used the Shapiro-Wilk test [34] to verify that our results do not exhibit normally distributed residuals, and Conover-Iman test for multiple comparisons [35]. We used the R package PMCMRplus [36]. Confidence intervals for the probability of outbreak were computed with Wilson’s method [37] implemented in the R package binom [38]. The model and all statistical analyses were implemented in R [39].

## Results

In the absence of interventions, the mean IFR is ∼ 2.5% in simulations where all severe cases requiring hospitalization recover (see Supplementary Fig. 1), and ∼ 11% in simulations where all severe cases die. We consider the latter scenario to evaluate the effect of non-medical preventive interventions. In this scenario, the probability of observing an outbreak is close to 0.84, in which ≿10% of the camp dies, the number of symptomatic cases peaks after 40 days, ∼84% of the population recovers, and ∼ 5% escape from infection (i.e. remain susceptible).

### Self-distancing

Our results show that self-distancing has a notable effect on reducing the probability of an outbreak which decreases roughly linearly as the percentage of contacts reduced increases. (see Fig. 3A). A near-quadratic trend is observed for the fraction of population dying, with a greater decrease for larger reduction in daily contacts, achieving a 34% reduction in mortality when the number of contacts is reduced by 50% (see Fig. 3B). Self-distancing also significantly extends the time until the peak of the outbreak, from ∼ 40 days when there is no intervention to 72 days when contacts are reduced by 50% (see Fig. 3C). The proportion of the population escaping from infection after 12 months also increases up to near 33% when a 50% reduction is considered, and the proportion of the population recovered after 12 months (which informs on the potential population level protection against future outbreaks) is reduced from 84% to 60% (see Supplementary Fig. 2).

**Figure 3:**
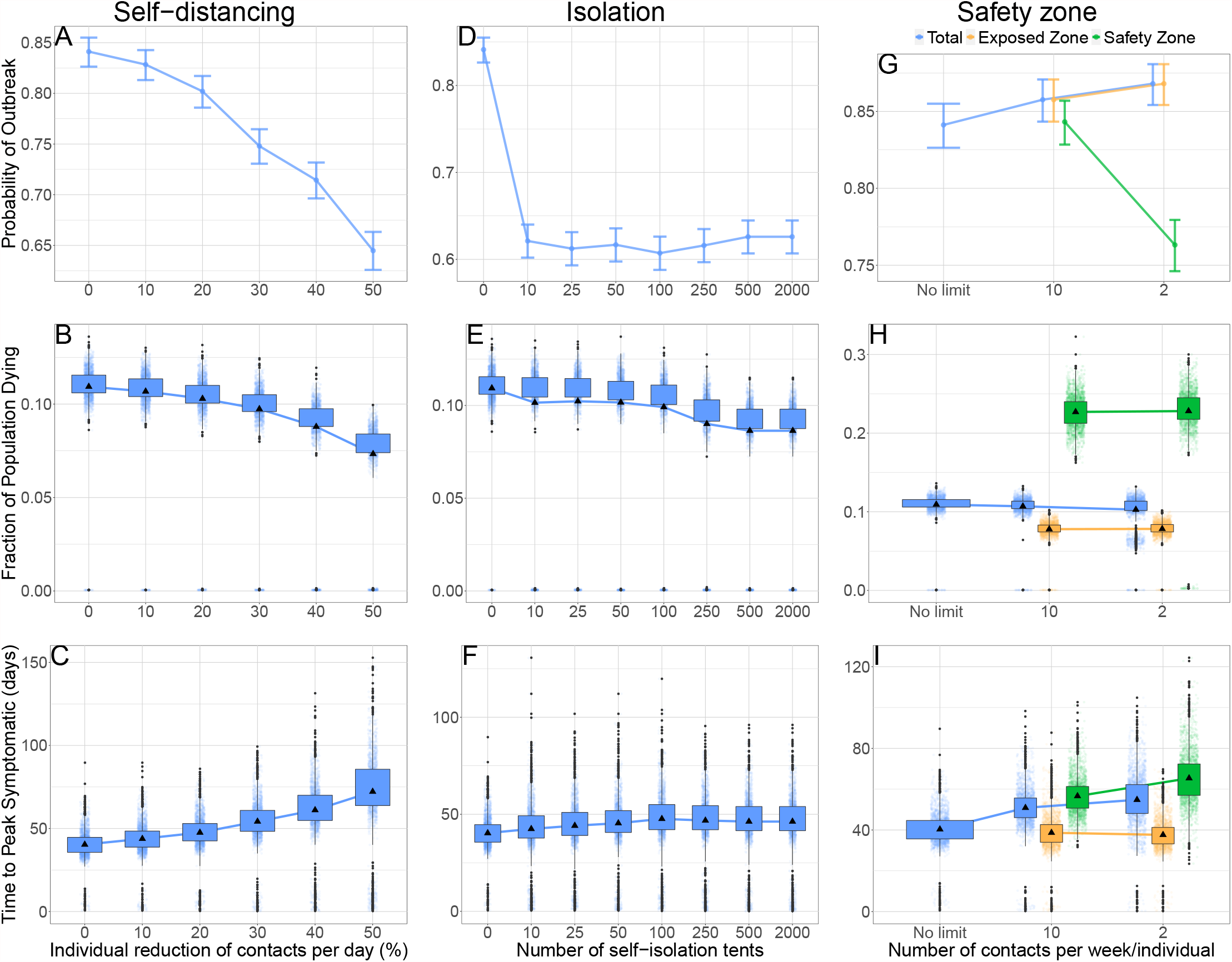
Effect of interventions on outbreak probability, fatalities and time until symptomatic cases peak. A: Self-distancing, probability of an outbreak. B: Self-distancing, fraction of the population dying. C: Self-distancing, time until peak symptomatic cases. D: Self-isolation, probability of an outbreak. E: Self-isolation, fraction of the population dying. F: Self-isolation, time until peak symptomatic cases. G: Safety zone, probability of an outbreak. H: Safety zone, fraction of the population dying. I: Safety zone, time until peak symptomatic cases. Triangles indicate the means and boxes interquartile ranges. Note that in figures of the safety zone intervention (panels G-I), the mean of an outcome for the whole population is not the weighted mean of the exposed and safety zones, since outcomes are computed considering simulations in which at least one death was observed in the population class inhabiting the zone, i.e. the number of simulations considered to compute each mean is different. In the safety zone figures (panels G-I) health-checks are in place, in Supplementary Fig. 5 we show the effect of removing health-checks.

### Self-isolation

With only 10 tents for a camp of 2000 people (i.e. 1 tent for every 200 people), self-isolation yields a marked decrease in the probability of outbreak (∼ 26%) (see Fig. 3D) but there is no significant reduction in the mortality (see Fig. 3E), suggesting that with a low number of tents the intervention is only effective in isolating index cases preventing the epidemic to start. It is needed to further increase the number of tents past this level up to at least 1 tent for every 8 people to observe a reduction of ∼ 18% in the mortality and an increase of ∼ 16% in the time to peak of symptomatic individuals. Increasing the number of tents does not further reduce the probability of outbreak. There is also an increase in the number of susceptible individuals at the end of the simulation with increasing number of tents. We finally observed an artificial increase in the IFR explained by an increasingly large number of simulations in which very few individuals are infected since, if at least one of them dies, we retrieve a high IFR value (see Suppl. Fig. 3).

### Safety zone

In this section, we consider the scenario in which all older adults, younger adults with comorbidities and their family members up to 20% of the camp population live in the green zone, unless otherwise specified. Creating a green zone improves the effect of the previous interventions overall, but with sometimes opposite outcomes in the exposed and protected populations. For example, the probability of an outbreak decreases for the protected population, by around 11%, if only two contacts are allowed per week in the buffer zone (see Fig. 3G). Notably, most of this reduction is only achieved when health-checks excluding symptomatic individuals from the buffer zone are in place (see Supplementary Fig. 5 for the effect of removing health-checks). On the other hand, the probability of an outbreak may slightly increase for the exposed population, a consequence of the relative increase in intra-zone contacts. By shifting the burden of an outbreak towards the less vulnerable population in the orange zone, another important outcome of this intervention is the notable increase in time (62%) until the number of symptomatic cases peaks for the vulnerable population, and a 40% increase in time for the whole population (see Fig. 3I). Nevertheless, this intervention only has a modest reduction on overall mortality (see Fig. 3H), possibly due to the high infectiousness of presymptomatic individuals.

Considering different scenarios for allocating people to the green zone, the lowest probability of an outbreak is achieved when only older adults or at most older adults and younger adults with comorbidities move there, with probabilities below 0.4 and 0.65, respectively (see Supplementary Fig. 8). Positive effects of the safety zone intervention are even more marked in camps with smaller populations, especially for probability of an outbreak in the green zone (which decreases by 55%) and overall mortality (which decreases by 20%) when the population is reduced from 2000 to 500. However, we also observe the adverse effect of a decrease in time until symptomatic cases peak (see Supplementary Fig. 9). The incorporation of a lockdown has the greatest effect on reducing the probability of an outbreak in the green zone, to under 0.25 when contacts in the buffer zone are reduced by 90%. While lockdowns show no positive effect on green zone fatalities in the few instances where an outbreak does reach there, they decrease overall IFR and fatalities by further concentrating outbreaks in the less vulnerable population (see Supplementary Fig. 10).

### Evacuation

We observe no significant effects when severe cases requiring hospitalization are evacuated (see Supplementary Fig. 4). Since we considered that these individuals will not receive health care (they are evacuated to isolation centers), their fate remains the same than if staying in the camp and, hence, we expect evacuation to have an effect only in reducing the infectivity. Although these individuals spend a longer period being infectious with respect to an individual having mild symptoms (∼ 10 days longer), the number of individuals under these conditions is only a small fraction of the total infectious population at any given time, which explains why we do not observe significant effects for this intervention.

### Combined interventions

The effects of the interventions observed when we examine them individually build upon each other when multiple interventions are implemented in tandem (see Fig. 4 and Supplementary Fig. 11). The protective effects of the safety zone intervention especially are most fully realized not when implemented on its own, but when paired with other interventions. They become so effective that outbreaks in the green zone become exceptionally rare, but so well controlled when they do happen, that the majority of outbreaks are small enough for us to observe an anomalous increase in IFR in some of the most effective interventions, driven by the discretization of the values it can take (e.g. if there is only one case and this person dies, see Supplementary Table. 11). When all interventions are implemented together: strict self-distancing (50% reduction in contacts), self-isolation of symptomatic cases (1 tent for every 40 people), a safety zone with 2 contacts per week in the buffer zone, health checks, a strict lockdown (90%), and evacuation of severe cases, mortality is reduced by ∼ 94% and the probability of outbreak in the green zone is very small (∼ 0.005). In other combinations with higher probability of outbreak (e.g. considering in the previous combination a 20% reduction in contacts instead of 50%) thetime to peak of symptomatic cases in the green zone is delayed by 48 days

**Figure 4:**
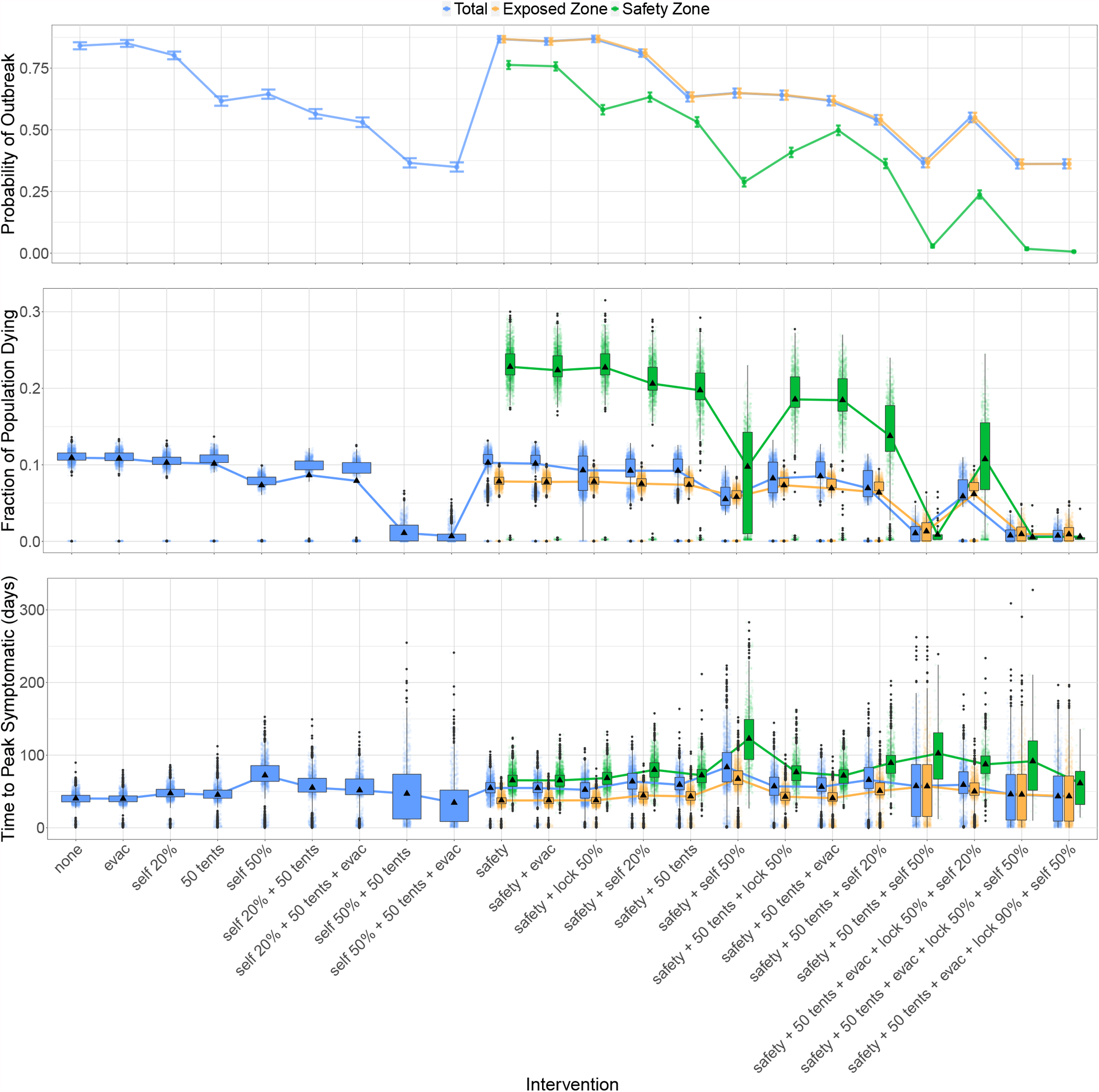
Combinations of interventions. Probability of an outbreak (top), fraction of the population dying (middle) and time until peak symptomatic cases (bottom) for different combination of interventions. Evac = evacuation of severely symptomatic, self = self-distancing, tents = number of available self-isolation tents, safety = safety zone, lock = lockdown of the buffer zone. For combinations of interventions including a safety zone, we distinguish between the population living in the green zone, in the orange zone and the whole population.

## Discussion

In this study, we propose a number of interventions of immediate applicability to informal settlements. We focused on IDP settlements in NW Syria, taking into account the interventions’ feasibility, cultural acceptance and their need for low-cost. When confronted with different possible scenarios, we generally considered the worst-cases, highlighting the interventions that are most effective in the direst conditions, but possibly resulting in an overestimate of mortality. This potential overestimation does not change the qualitative picture of the results, which is built upon comparison of relative values between the presence and absence of interventions.

Our results align with previous simulation studies of potential COVID-19 interventions in similarly densely populated, low-resource settings where informal settlements are present, such as urban areas of sub-Saharan Africa. In these settings, social-distancing is demonstrated to be an effective intervention, and even small changes are estimated to have large effects on outbreaks [40], in some cases determining whether or not already inadequate healthcare systems become overwhelmed [41]. Zandvoort et al. show that similar measures to the ones we consider: self-isolation, physical distancing and “shielding” the vulnerable, may reduce mortality by 60%-75% in African cities [12].

Self-distancing proves to be an effective measure in our models as well; reducing contacts by 50% has the greatest effect across most outcomes of interest in any of the interventions we examined. However, the difficulty of achieving a reduction of this magnitude cannot be overlooked, especially considering the large proportion of the population composed of children, a group with an already high contact rate that may prove difficult to control [23]. To illustrate the micro-dynamics of this intervention, in Supplementary Fig. 12 we plot the maximum proportion of the population exposed at any given point in time in each simulation, against the time it takes for symptomatic cases to peak. We observe that until reaching approximately 60%, successive reductions in contacts reduce the maximum proportion of the population exposed while increasing the time until symptomatic cases peak. Additional reductions in contacts beyond 60% however abruptly decrease both the time until cases peak and overall mortality, suggesting that outbreaks die out before the virus spreads widely throughout the camp’s population. This suggests the existence of a critical threshold for the number of individuals exposed over which large outbreaks become established in the population, significantly increasing mortality.

We also propose self-isolation using individual tents which can be located in a dedicated zone or next to the tents of relatives, where contact with non-isolated individuals is mediated by a buffer zone. This intervention is effective with even a small number of isolation tents, as low as 5-10 tents per 1000 camp residents in preventing an outbreak in the camp, but it requires at least 125 tents per 1000 camp residents to substantially reduce mortality After conversations with camp managers, we found that this intervention is more likely to be accepted in NW Syria than evacuation to community-based isolation centers. Community-based isolation not only poses cultural challenges; the capacity required to implement it has hardly been met [30], and it is still one of the main challenges in the region [8]. We note that in our simulated intervention individuals become isolated as soon as they have symptoms. Recognizing symptoms, however, may require some time and we should expect a less effective intervention unless systematic checks for symptomatic individuals are put in place.

Setting up a safety zone has two positive effects that most stand out: a reduction in the probability of an outbreak in the vulnerable population, and an increase in the time until the number of symptomatic cases peaks. Much of the success or failure of the safety zone intervention hinges on the functioning of the buffer zone. The number of inter-zone contacts per week, the implementation of health checks, and potential lockdowns all have notable effects. Also important is the portion of the population that is protected; protecting only the vulnerable may have the most beneficial effects, but it is precisely these vulnerable individuals, older adults and people with comorbidities, who may most need family members to care for them. While safety zone scenarios that allow greater numbers of family members to accompany their vulnerable relatives to the green zone may confer greater epidemiological risk, they may also engender greater well-being and social cohesion.

Despite these benefits, we do not observe a clear decrease in IFR with this intervention, although it is possible that our model may overestimate mortality from an outbreak in the green zone in the few instances when there is one. Since it is unlikely that the camps have the economic means to increase the number of tents when implementing this intervention, we assumed that individuals do not reduce their contacts when moved to the green zone since household sizes will not decrease, which implies an increase in the number of contacts between vulnerable individuals. Despite this increase in contacts, we do not observe an increase in mortality in the vulnerable population when the safety zone is implemented. These results address concerns raised around this type of intervention from previous experiences with large numbers of fatalities registered in nursing-homes in developed countries [42]. While nursing-homes in developed countries may be seen as analogous to the safety zone intervention, the alternative to nursing homes in developed countries for the elderly population typically involves remaining at home with few contacts with younger individuals (in a scenario of lockdown), while in the camps the alternative is living in tents shared with younger individuals with high contacts rates (especially children). This may explain why we observe a positive effect from this intervention despite a relative increase in contacts among the more vulnerable subpopulation.

An instrumental consideration for our models is the fraction of the population recovered from COVID-19 after a steady state is reached. Although the duration for which SARS-CoV-2 infection confers immunity is uncertain, the proportion of the population recovered after an outbreak should play a role in its protection against future ones. For all intervention except self-distancing *>*30%, we observed that the fraction of the population recovered meets or exceeds 50%.

Another important consideration for interpreting our results are the modelling assumptions we made. From the point of view of our parameterization, perhaps the most relevant relates to the relative transmissivity of the different infectious stages, whose specific values still have large margins of variability [43, 44, 28]. This is particularly relevant for our purposes, because the higher the relative infectivity of the presymptomatic and asymptomatic stages, the less effective our non-medical interventions which rely on identification of symptomatic individuals will be. For instance, Bullock et al. [45] assumed a higher infectiousness of the presymptomatic stage and hence self-isolation of symptomatic individuals had little effect. Self-isolation also becomes ineffective under the assumptions made by Hernandez-Suarez et al. [46] when they considered isolating only severe symptomatic cases (whose fraction is small), hence mildly symptomatic individuals were effectively considered asymptomatic. On the other hand, Gilman et al. [47] showed that self-isolation was effective when considering individuals at different stages to be equally infectious. We assumed that presymptomatic individuals have the highest relative infectivity, consistent with the most recent estimations [44], and the interventions we proposed are still effective. Considering a different scenario in which all compartments are assumed to be equally infectious, our interventions become even more effective (see previous version of our manuscript [48]v1).

It is also important to acknowledge the benefits and limitations of different possible computational implementations. For instance, there are interventions that do not have a natural implementation within our framework, such as those requiring interventions targeting very specific interactions between individuals (as opposed to large groups of individuals), or reproducing empirically-observed residence times. An example might be the isolation of an individual and his/her family, as proposed by Gilman *et al*. [47] which, since we do not explicitly model interactions at the family-level, would require the creation of as many classes as families. When this level of detail is required, individual based models (IBMs) may be more appropriate [47, 45]. However, IBMs require a rich amount of data for their parameterization which, although increasingly available, is scarce for informal IDPs camps. Our framework is powerful enough to simulate a large number of scenarios with little computational cost, which would be an optimal strategy as a first approximation in the design of interventions to narrow down the most relevant scenarios (as a reference, in *<*24h we model with just 12 cores 75 scenarios requiring quarter million simulations). The scenarios selected could then be further investigated with more detailed interventions using IBMs, if data is available.

A key limitation of our approach is that it simulates an outbreak started by one infectious individual in a single camp with a closed population. We acknowledge that this approach does not fully capture the complexities of the NWS region, where IDPs live interspersed throughout the region in several hundred camps. The dynamics of an outbreak in the region are undoubtedly influenced by inter-community contacts, and the dynamics of an outbreak in a single camp by these region-wide dynamics, as it has been demonstrated in other countries [10, 49]. We expect our results to be robust to changes in population, as long as these changes are relatively small compared to the total population size in the camp, implying sporadic inputs of infected individuals. This is the expected behaviour in IDPs, which are often small and located in rural areas, and in which important population movements, as those observed in large camps, are infrequent. This fact, together with the relatively young population in IDPs, may help in limiting the impact of the disease, as observed in African rural areas [50].

Other unaccounted for social and cultural dynamics will undeniably complicate the feasibility of our proposed interventions. Only one example we have not addressed here is the unlikeliness of children under 13 self-isolating. Although the number of challenges to implementing our proposed interventions are potentially endless, the community-based nature of our approach may help circumvent these challenges much faster than healthcare-based interventions, which often depend on complex political decisions and may take years to build the requisite capacity for an effective response. If the dynamics of the virus are well understood by local communities and at least some of the interventions we propose are implemented, the impacts of COVID-19 can be mitigated even in an environment as challenging as NW Syria.

## Conclusion

Given a rapidly changing environment and slow responses of local and international authorities in conflict regions where control of policy is disputed, the latter often leaving these communities aside in their priorities [51], empowering local communities themselves is perhaps the best, if not the only way, to help them avoid the worst consequences of the pandemic. Such an approach may achieve greater compliance with non-pharmaceutical interventions, especially where there is a mistrust of external authority. This not only applies to IDP camps in NW Syria, but more generally to refugee camps in conflict-torn regions, and potentially other informal settlements and vulnerable communities around the world: the low-cost, effective interventions we present are feasible, needed and urgent.

## Data Availability

The data used in this study was publicly available

https://github.com/crowdfightcovid19/req-550-Syria

## Acknowledgements

This collaboration was organized by crowdfightCOVID19 (www.crowdfightcovid19.org) upon request from CS. We thank Judith Boumann for valuable contributions. We thank Peter Ashcroft, Juan Poyatos, Noreen Goldman, Burcu Tepekule and members of Sebastian Bonhoeffer’s and Bryan Grenfell’s groups for useful discussions. We thank two anonymous reviewers who provided thoughtful and insightful comments that greatly improved our article.

## Declarations

### Ethics approval

This study used only publicly available aggregate data and was thus not subject to ethical review.

### Consent to participate

NA

### Consent for publication

All authors agreed on publication.

### Availability of data and material

All results are available at the url https://github.com/crowdfightcovid19/req-550-Syria

### Code availability

All the code is freely available at the url https://github.com/crowdfightcovid19/req-550-Syria

## Authors’ contributions

All authors contributed to the conceptualization. Design of the methodology: APG, ECF, JV, JK, CS. Formal analysis: APG, ECF, JK. Code development APG, ECF, JK. Conducted research: APG, ECF, JV, JK. Validate results: APG, ECF, JK, JV, CS. Contributed resources: APG, CS, ECF, JK, JV. Data curation: APG, JK, ECF. Visualization: APG, ECF, JK, JV. Writing (original draft) APG, ECF, JK. All authors contributed to the final version of the manuscript, and APG supervised the research.

## Abbreviations

IDP: Internally Displaced Persons
NWS: Northwest region of Syria
IFR: Infection fatality rate
ICU: Intensive care unit
IBM: Individual based models

