## Supplementary Materials for "Empowering the crowd: Feasible strategies for epidemic management in high-density informal settlements. The case of COVID-19 in Northwest Syria"

July 1, 2021

(1) Institute of Integrative Biology. ETH-Zürich. Zürich, Switzerland.
(2) Office of Population Research. Princeton University. Princeton, NJ, USA.
(3) Princeton Environmental Institute. Princeton University. Princeton, NJ, USA.
(4) Genome Damage and Stability Centre. University of Sussex. Brighton, United Kingdom.
(5) Pax Syriana Foundation. Valetta, Malta.
(∧) Equal contribution.

### 1 Parameterization of the model

#### 1.1 Derivation of fixed parameters (Table 1 in Main Text)

To estimate the latent period ($1/\delta_{\mathrm{E}}$), we calculated the difference between randomly generated incubation ($1/\delta_{\mathrm{E}}+1/\delta_{\mathrm{P}}$) and presymptomatic ($1/\delta_{\mathrm{P}}$) periods. We estimated the presymptomatic period using results reported by He et al. [1] and found they best fit a Gompertz distribution with a mean of 2.3 days (95% CI: 0.8-3.0). Since a correction of these by Ashcroft et al. [2] suggests they significantly underestimate the presymptomatic period's upper bound, we estimated that the true presymptomatic period should rather be closer to a Gaussian distribution around the mean (95% CI: 0.8-3.8). However, this presymptomatic period distribution implies a non-negligible probability of a negative latent period. To correct this discrepancy, we assumed a minimum latent period of .5 days [3]. Time from symptom onset to death in critical cases ($1/\alpha$) is estimated using time from symptom onset to ICU admission in Wang et al [4].

#### 1.2 Population structure of demographic-classes derivation (Table 2 in Main Text)

In April, 2020, 40.7% of the population in informal IDP camps in Northern Syria was aged 0-12, 53.4% aged 13-50, and 5.9% aged 51+ [5]. To estimate the proportion of each age group with comorbidities, we calculated the weighted average age-specific comorbidity prevalence of the 4 most common comorbidities in the Syrian refugee populations in Jordan and Lebanon: hypertension, cardiovascular disease, diabetes, and chronic respiratory disease [6, 7]. We standardized these weighted averages to the age structure of IDPs in Northern Syria and estimated that 11.7% of people aged 13-50 have comorbidities, while 62.9% of people aged 51+ have comorbidities.

#### 1.3 Derivation of transmissibility parameters

The probability of infection if there is a contact between a susceptible and an infected person depends on the stage of the disease, denoted $\tau\beta_{\mathrm{P}}$, $\tau\beta_{\mathrm{A}}$, $\tau\beta_{\mathrm{I}}$ or $\tau\beta_{\mathrm{H}}$ depending upon whether the infected individual is in the presymptomatic ($P_i$), symptomatic ($I_i$), asymptomatic ($A_i$), or hospitalized compartment ($H_i$), respectively. We estimated these parameters in two steps. In the following section, we estimate the $\beta_{\mathrm{X}}$ parameters ($\mathrm{X} \in \{\mathrm{P}, \mathrm{A}, \mathrm{I}, \mathrm{H}\}$) which represent the relative transmissibility of each stage with respect to the maximum transmissibility $\tau$. After this calculation, we present our estimate for the maximum transmissibility parameter $\tau$.

###### Relative transmissibilities $\beta$ (Table 1 in Main Text)

We start by considering the transmissibility of the presymptomatic stage for those individuals who become symptomatic as a reference ($\beta_{\mathrm{P}\to\mathrm{I}} = 1$), since the probability of infection from contact with an individual at this epidemiological stage is highest [8]. Next, we set the contribution of each epidemiological stage to infectivity as proportional to $\beta_{\mathrm{X}}/\gamma_{\mathrm{X}}$, with $1/\gamma_{\mathrm{X}}$ the duration of stage X. Following He et al, we estimate the proportion of infectivity in individuals who go on to develop symptoms that occurs at the presymptomatic stage ($\mathrm{X} \in \{\mathrm{P}\}$) as the area under the infectivity curve prior to symptom onset, $AUC_{\mathrm{P}}$, and the proportion of infectivity that occurs at symptomatic stages ($\mathrm{X} \in \{\mathrm{I}, \mathrm{H}\}$) as the area under the infectivity curve after symptom onset, $1 - AUC_{\mathrm{P}}$ [8]:

$$\frac{AUC_{\mathrm{P}}}{(1 - AUC_{\mathrm{P}})} \approx \frac{\frac{\beta_{\mathrm{P}\to\mathrm{I}}}{\delta_{\mathrm{P}}}}{\frac{\beta_{\mathrm{I}}}{\gamma_{\mathrm{I}}} + \frac{\beta_{\mathrm{H}}}{\gamma_{\mathrm{H}}}} \tag{1}$$

We then considered the quantity $\rho_{\mathrm{HI}}$, the ratio of the viral culture positive test rate in hospitalized patients 7-16 days since start of symptoms to the positive test rate in patients 0-6 days since start of symptoms from van Kampen et al.[9]. Similarly, the relative risk of asymptomatic transmission to symptomatic transmission according to Byambasuren et al. is expressed as $\rho_{\mathrm{AI}}$ [10]:

$$\beta_{\mathrm{A}} = \rho_{\mathrm{AI}}\beta_{\mathrm{I}} \tag{2}$$

$$\beta_{\mathrm{H}} = \rho_{\mathrm{HI}}\beta_{\mathrm{I}} \tag{3}$$

Considering these relationships we rewrite Eq. 1 to obtain the desired parameters:

$$\beta_{\mathrm{I}} = \frac{\beta_{\mathrm{P}\to\mathrm{I}}\gamma_{\mathrm{I}}\gamma_{\mathrm{H}}(1 - AUC_{\mathrm{P}})}{AUC_{\mathrm{P}}\delta_{\mathrm{P}}(\gamma_{\mathrm{H}} + \rho_{\mathrm{HI}}\gamma_{\mathrm{I}})} \quad (4)$$

$$\beta_{\mathrm{A}} = \frac{\rho_{\mathrm{AI}}\beta_{\mathrm{P}\to\mathrm{I}}\gamma_{\mathrm{I}}\gamma_{\mathrm{H}}(1 - AUC_{\mathrm{P}})}{AUC_{\mathrm{P}}\delta_{\mathrm{P}}(\gamma_{\mathrm{H}} + \rho_{\mathrm{HI}}\gamma_{\mathrm{I}})} \quad (5)$$

$$\beta_{\mathrm{H}} = \frac{\rho_{\mathrm{HI}}\beta_{\mathrm{P}\to\mathrm{I}}\gamma_{\mathrm{I}}\gamma_{\mathrm{H}}(1 - AUC_{\mathrm{P}})}{AUC_{\mathrm{P}}\delta_{\mathrm{P}}(\gamma_{\mathrm{H}} + \rho_{\mathrm{HI}}\gamma_{\mathrm{I}})}. \quad (6)$$

The values of $AUC_{\mathrm{P}}$, $\rho_{\mathrm{AI}}$ and $\rho_{\mathrm{HI}}$ are presented in Table 1, and the values of $\beta_{\mathrm{X}}$ in Main Text. Noting that the starting point for these derivations is the transmissibility of individuals in the presymptomatic stage who go on to develop symptoms ($\beta_{\mathrm{P}\to\mathrm{I}} = 1$), we need to go back and derive the transmissibility of all presymptomatic individuals ($\beta_{\mathrm{P}}$), taking those that will be asymptomatic into account. We assumed that the relative transmissibility of presymptomatic individuals who will become asymptomatic to those that will become symptomatic, is equal to the relative transmissibility of asymptomatic to symptomatic individuals ($\rho_{\mathrm{AI}}$). With the proportion of presymptomatic cases that will become developed symptoms $f$ (encoded in in Eqs. 1–8 in Main Text), we compute the mean transmissibility of all presymptomatic individuals as:

$$\beta_{\mathrm{P}} = f\beta_{\mathrm{P}\to\mathrm{I}} + (1 - f)\rho_{\mathrm{AI}}\beta_{\mathrm{P}\to\mathrm{I}}. \quad (7)$$

| Parameter | Description | Value | Distribution | Reference |
|---|---|---|---|---|
| $AUC_{\mathrm{P}}$ | Presymptomatic area under infectivity curve | 0.44 (95% CI: .30-.57) | Gaussian | [8] |
| $\rho_{\mathrm{AI}}$ | Ratio of asymptomatic to symptomatic infectiousness | 0.58 (95% CI: .34-.99) | Lognormal | [10] |
| $\rho_{\mathrm{HI}}$ | Ratio of hospitalized to symptomatic infectiousness | 0.48 | - | [9] |

Table 1: **Relative transmissibility parameters.**

##### Maximum transmissibility parameter $\tau$

In the following, to simplify the notation we define $\kappa_i = (l_i\gamma_{\mathrm{I}} + h_i\eta + g_i\alpha)$. To estimate the probability of infection if there is a contact between a susceptible and an infected individual (parameter $\tau$) we proceed as follows [11, 12, 13]. We start by considering the subsystem containing the infected population:

$$\dot{E}_i = \lambda_i S_i - \delta_{\mathrm{E}} E_i \quad (8)$$

$$\dot{P}_i = \delta_{\mathrm{E}} E_i - \delta_{\mathrm{P}} P_i \quad (9)$$

$$\dot{A}_i = (1 - f)\delta_{\mathrm{P}} P_i - \gamma_{\mathrm{A}} A_i \quad (10)$$

$$\dot{I}_i = f\delta_{\mathrm{P}} P_i - \kappa_i I_i \quad (11)$$

$$\dot{H}_i = h_i\eta I_i - \gamma_{\mathrm{H}} H_i. \quad (12)$$

For the sake of simplifying the notation, let us consider the following ordering of the variables in the vector $x = (E_1, ..., E_{\mathrm{M}}, P_1, ..., P_{\mathrm{M}}, A_1, ..., A_{\mathrm{M}}, I_1, ..., I_{\mathrm{M}}, H_1, ..., H_{\mathrm{M}})$, with $M$ the number of population classes. We are interested in the parameterization of the null model, which will serve as a baseline to estimate the parameter $\tau$, which is initially unknown, but does not change when interventions are introduced. Considering the contacts matrix for the null model (Eq. 9 in Main Text), the rate of exposure becomes

$$\lambda_i = \frac{\tau}{N}\sum_{j=1}^{M} c_i\left(\beta_{\mathrm{P}} P_j + \beta_{\mathrm{A}} A_j + \beta_{\mathrm{I}} I_j + \beta_{\mathrm{H}} H_j\right). \quad (13)$$

In the following, we use bold symbols for vectors and matrices, and the symbols $\odot$ and $\oslash$ for the element-wise multiplication and division, respectively. Following this notation, the linearized system can be written in the form $\dot{\boldsymbol{x}} = (\boldsymbol{T} + \boldsymbol{\Sigma})\boldsymbol{x}$, where:

$$\boldsymbol{T} = \tau \begin{bmatrix} \mathbf{0} & \boldsymbol{\Theta}_\text{P} & \boldsymbol{\Theta}_\text{A} & \boldsymbol{\Theta}_\text{I} & \boldsymbol{\Theta}_\text{H} \\ \mathbf{0} & \mathbf{0} & \mathbf{0} & \mathbf{0} & \mathbf{0} \\ \mathbf{0} & \mathbf{0} & \mathbf{0} & \mathbf{0} & \mathbf{0} \\ \mathbf{0} & \mathbf{0} & \mathbf{0} & \mathbf{0} & \mathbf{0} \\ \mathbf{0} & \mathbf{0} & \mathbf{0} & \mathbf{0} & \mathbf{0} \end{bmatrix} \tag{14}$$

is the transmission matrix, with $\boldsymbol{\Theta}_\text{X} = \beta_\text{X}\text{diag}(\boldsymbol{p} \odot \boldsymbol{c})\boldsymbol{U}$, $\boldsymbol{p} = \boldsymbol{N}/N$, $\boldsymbol{U}$ is the all-ones matrix of size $M$, and $\beta_\text{X}$ the infectiousness of compartment X relative to the presymptomatic compartment (see Main Text for details). The transition matrix is

$$\boldsymbol{\Sigma} = \begin{bmatrix} -\delta_\text{E}\boldsymbol{I} & \mathbf{0} & \mathbf{0} & \mathbf{0} & \mathbf{0} \\ \delta_\text{E}\boldsymbol{I} & -\delta_\text{P}\boldsymbol{I} & \mathbf{0} & \mathbf{0} & \mathbf{0} \\ \mathbf{0} & (1-f)\delta_\text{P}\boldsymbol{I} & -\gamma_\text{A}\boldsymbol{I} & \mathbf{0} & \mathbf{0} \\ \mathbf{0} & f\delta_\text{P}\boldsymbol{I} & \mathbf{0} & -\text{diag}(\boldsymbol{\kappa})\boldsymbol{I} & \mathbf{0} \\ \mathbf{0} & \mathbf{0} & \mathbf{0} & \eta\text{diag}(\boldsymbol{h})\boldsymbol{I} & -\gamma_\text{H}\boldsymbol{I} \end{bmatrix} \tag{15}$$

Where $\boldsymbol{I}$ and $\mathbf{0}$ are the identity and null matrices of size $M$, and $\boldsymbol{\kappa} = \boldsymbol{l}\gamma_I + \boldsymbol{h}\eta + \boldsymbol{g}\alpha$. We next compute the inverse of the transition matrix

$$\boldsymbol{\Sigma^{-1}} = \begin{bmatrix} -\frac{1}{\delta_\text{E}}\boldsymbol{I} & \mathbf{0} & \mathbf{0} & \mathbf{0} & \mathbf{0} \\ -\frac{1}{\delta_\text{P}}\boldsymbol{I} & -\frac{1}{\delta_\text{P}}\boldsymbol{I} & \mathbf{0} & \mathbf{0} & \mathbf{0} \\ -\frac{(1-f)}{\gamma_\text{A}}\boldsymbol{I} & -\frac{(1-f)}{\gamma_\text{A}}\boldsymbol{I} & -\frac{1}{\gamma_\text{A}}\boldsymbol{I} & \mathbf{0} & \mathbf{0} \\ -f\text{diag}(\boldsymbol{\kappa}^{-1})\boldsymbol{I} & -f\text{diag}(\boldsymbol{\kappa}^{-1})\boldsymbol{I} & \mathbf{0} & -\text{diag}(\boldsymbol{\kappa}^{-1})\boldsymbol{I} & \mathbf{0} \\ -\frac{f\eta}{\gamma_\text{H}}\text{diag}(\boldsymbol{h} \oslash \boldsymbol{\kappa})\boldsymbol{I} & -\frac{f\eta}{\gamma_\text{H}}\text{diag}(\boldsymbol{h} \oslash \boldsymbol{\kappa})\boldsymbol{I} & \mathbf{0} & -\frac{\eta}{\gamma_\text{H}}\text{diag}(\boldsymbol{h} \oslash \boldsymbol{\kappa})\boldsymbol{I} & -\frac{1}{\gamma_\text{H}}\boldsymbol{I} \end{bmatrix} \tag{16}$$

The NGM with large domain can now be found by $\boldsymbol{K_\text{L}} = -\boldsymbol{T}\boldsymbol{\Sigma}^{-1}$. However, since we know that each individual who gets infected becomes exposed ($E$ compartment), we focus on the NGM with small domain, $\mathbf{K}_\text{S}$, which only consists of the $E$ compartment [14]. We do this by removing the rows that correspond to the other compartments from $T$ and the columns from $\Sigma^{-1}$ . We then find:

$$\boldsymbol{K_\text{S}} = \tau \left[ \frac{1}{\delta_P}\boldsymbol{\Theta}_\text{P} + \frac{(1-f)}{\gamma_A}\boldsymbol{\Theta}_\text{A} + f\text{diag}\left(\boldsymbol{h}^{-1}\right)\boldsymbol{\Theta}_\text{I} + \frac{f\eta}{\gamma_\text{H}}\text{diag}(\boldsymbol{h} \oslash \boldsymbol{\kappa})\boldsymbol{\Theta}_\text{H} \right]. \tag{17}$$

The reproduction number is related to the dominant eigenvalue of $\boldsymbol{K_\text{S}}$, i.e. $R_0 = |\lambda_1|$, and $\tau$ is estimated from the real dominant eigenvalue of $\tilde{K}_\text{S} = K_\text{S}/\tau$. Considering the null model parameters ($\tilde{\lambda}_1^0$), we have the expression:

$$\tau = \frac{R_0}{|\tilde{\lambda}_1^0|}. \tag{18}$$

#### 1.4 Epidemiological severity proportions (Table 4 in Main Text)

In the Main Text, we presented the proportions in which clinical symptomatic individuals resolve into critical ($q_i^D$), severe ($q_i^\text{H}$) and recovered ($q_i^R$) cases. We assigned the fractions of symptomatic cases in children aged <13 that would become severe and critical from the fractions of symptomatic cases in children aged <11 that were severe and critical in China [15]. We assigned the class-specific fractions of symptomatic cases in adults that would become severe and critical based on age and comorbidity-specific fractions of symptomatic cases with known outcomes that required hospitalization, without and with ICU admission, respectively in the United States [16]. To account for poorer health among Syrian adults compared to their similarly aged peers in developed countries, estimates for US adults aged 19-64 were used for Syrian adults aged 13-50, while estimates for US adults aged 65+ were used for Syrian adults aged 51+.

Since the rates at which these individuals progress are different ($\eta$ for $H$, $\alpha$ for $D$ and $\gamma_{\mathrm{I}}$ for $R$) we introduced three parameters, $h_i$, $g_i$ and $l_i$, to distribute individuals according to the desired proportions following the equations:

$$q_i^{\mathrm{H}} = h_i \eta \kappa_i^{-1} \tag{19}$$

$$q_i^{D} = g_i \alpha \kappa_i^{-1} \tag{20}$$

$$q_i^{R} = 1 - q_i^{\mathrm{H}} - q_i^{D} = l_i \gamma_{\mathrm{I}} \kappa_i^{-1}, \tag{21}$$

where $\kappa_i = h_i\eta + g_i\alpha + l_i\gamma_{\mathrm{I}}$. The system has three unknowns and three equations but one equation linearly depends on the other two, hence we introduce the constraint $l_i = 1 - h_i - g_i$ to solve the system as:

$$h_i = \frac{\alpha q_i^H}{\eta q_i^D} g_i, \tag{22}$$

$$g_i = \gamma_I \left( \frac{\alpha}{q_i^D} + \frac{\gamma_I \alpha q_i^H}{\eta q_i^D} + \gamma_I - \frac{\alpha q_i^H}{q_i^D} - \alpha \right)^{-1}. \tag{23}$$

### 2 Parameterization of the interventions (Table 5 in Main Text)

#### 2.1 Safety zone

We considered the existence of a safety zone to protect a certain fraction, $f_{\mathrm{S}}$, of the population, mostly those more vulnerable. In practice, this involves dividing the camp in two areas, a "green" zone (denoted g) for the protected population and an "orange" zone (o) for the exposed population, and dividing each demographic-class into two behaviour-classes for each respective zone. These two populations interact via a buffer zone, under controlled conditions where we assumed transmissivity is reduced by 80%, encoded in the parameter $\xi_{ij} = 0.2$. Each individual in the green zone can interact with a limited number ($c_{\mathrm{visit}}$) of family members (hereafter "visitors") from the orange zone per day. In some interventions we considered that individuals visiting the buffer zone will have a health check (e.g. temperature measurement), aimed at excluding symptomatic individuals. When the health check is applied, the probability of transmission by individuals in the $I$ or $H$ compartments from one zone to susceptible individuals from a different zone is set to zero (see parameters $\zeta_{\mathrm{I}}$ and $\zeta_{\mathrm{H}}$ in Eq. 24 in Main Text). In the following, we derive the values of parameters $\epsilon_{ij}$ and $\omega_{ij}$, modifying the rate at which individuals become exposed (see Eq. 24 in Main Text).

Although setting up a safety zone implies a reduction in the number of contacts between classes of the green zone and the orange zone, the mean number of contacts that each individual has per day, $c_i$, is conserved. Therefore we need to estimate how contacts will be redistributed from individuals from a different zone to individuals living in the same zone. We model this redistribution of contacts with the parameter $\epsilon_{ij}$:

$$\begin{aligned} \epsilon_{ij} &= \vartheta c_{\mathrm{visit}}/c_i \quad (i, j \text{ in different zones}) \\ \epsilon_{ij} &= 1 - \vartheta c_{\mathrm{visit}}/c_i \quad (i, j \text{ in same zone}). \end{aligned}$$

We define $\vartheta$ as[1]:

$$\vartheta = \begin{cases} 1 & \text{if } i \in \mathrm{g} \\ f_{\mathrm{o,visit}} & \text{if } i \in \mathrm{o} \end{cases}$$

[1]If $c_{\mathrm{visit}}$ is large enough ( $c_{\mathrm{visit}} \approx 15$ contacts per day), $\vartheta$ should saturate, because every member of the orange zone would eventually visit the buffer zone, following the expression:

$$\vartheta = \begin{cases} 1 & \text{if } i \in \mathrm{g} \\ f_{\mathrm{o,visit}} \left(1 - \Theta(f_{\mathrm{o,visit}} - 1)\frac{f_{\mathrm{o,visit}}-1}{f_{\mathrm{o,visit}}}\right) & \text{if } i \in \mathrm{o} \end{cases}$$

with the Heaviside function $\Theta(f_{\mathrm{o,visit}} - 1) = 1$ if $f_{\mathrm{o,visit}} \geq 1$. We chose values well below this saturation threshold (a maximum of 10 contacts per week, i.e. 1.42 contacts per day).

If we assume that visitors are always different, the quantity $f_{\rm o,visit} = c_{\rm visit}\frac{N_{\rm g}}{N_{\rm o}}$ is the fraction of the orange population that visits the buffer zone.

Next, we estimate how the probability of interaction between a member of class $i$ and class $j$ is modified with respect to the null model, depending on the zones from which class $i$ and class $j$ are drawn. Suppose classes $i$ and $j$ are separated from the rest of the camp's population and confined within a restricted zone. The probability that an individual of class $i$ randomly encounters an individual of class $j$ would now be higher than in a well-mixed population where interaction with individuals belonging to other classes is not restricted. This modification of relative probability of interaction is encoded in the parameter $\omega_{ij}$ (see Eq. 24 in Main Text). More specifically, the proportion $N_i/N$ of individuals of class $i$ in the null model becomes $N_i/N_{\rm X}$ with $N_{\rm X}$ the total number of individuals in zone $X = \{\rm o, g\}$. This yields the following values for $\omega_{ij}$:

$$\begin{aligned}
\omega_{ij} &= \left(\tfrac{N_i}{N_{\rm X}}\right)/\left(\tfrac{N}{N_i}\right) = \frac{N}{N_{\rm X}} \quad (i, j \text{ in same zone } X) \\
\omega_{ij} &= \left(\tfrac{N_i}{N_{\rm Y}}\right)/\left(\tfrac{N}{N_i}\right) = \frac{N}{N_{\rm Y}} \quad (i \in X \text{ and } j \in Y).
\end{aligned}$$

| Scenario | Age 1, orange | Age 1, green | Age 2 no comorbidities, orange | Age 2 no comorbidities, green | Age 2 comorbidities, orange | Age 2 comorbidities, green | Age 3 no comorbidities, green | Age 3 comorbidities, green |
|---|---|---|---|---|---|---|---|---|
| Only age 3 in green zone | .407 | 0 | .471 | 0 | .0626 | 0 | .022 | .0373 |
| Age 3 + age 2 with comorbidities in green zone | .407 | 0 | .471 | 0 | 0 | .0626 | .022 | .0373 |
| 20% green zone capacity | .376 | .0312 | .424 | .0469 | 0 | .0626 | .022 | .0373 |
| 25% green zone capacity | .356 | .0512 | .394 | .0769 | 0 | .0626 | .022 | .0373 |
| 30% green zone capacity | .336 | .0712 | .364 | .107 | 0 | .0626 | .022 | .0373 |

Table 2: **Fraction of population in each zone by safety zone scenario and behaviour-class.** Behaviour-classes that are not considered in a given scenario have a proportion equal to zero.

Following this parameterization, we explore different scenarios, summarized in Table 2, for allocating members of each population class to the safety, or "green" zone, and the exposed, or "orange" zone. In one scenario, we only place individuals in age group 3 (>50) in the green zone, while in another we place all vulnerable individuals, age group 3 and age group 2 (13-50) with comorbidities, in the green zone. In 3 additional scenarios, after all vulnerable individuals are allocated to the green zone, we set the green zone's capacity to a certain percentage of the camp's population (20%, 25%, 30%), and allocate its remainder to non-vulnerable family members, who by necessity are either children <13 in age group 1 or healthy younger adults in age group 2. In accordance with camp managers' expectations that many vulnerable individuals will have non-vulnerable spouses, while fewer vulnerable individuals will have young children, in these scenarios we allocate 40% of the remainder of the green zone to children and 60% of the remainder of the green zone to younger adults without comorbidities. We also consider a baseline scenario in which there is no green zone.

#### 2.2 Self-isolation and evacuation

To implement self-isolation it is required a delineation between the isolated and non-isolated population within the clinical symptomatic compartment after identification of symptoms ($I_i$). This does not entail the creation of a new compartment, but rather the estimation of the number of individuals in each class in isolation ($\tilde{I}_i$). We first compute the total number of infected individuals across every class, $N_{\mathrm{I}} = \sum_i I_i$, at each integration step in the simulation. Next, we consider the isolation capacity of the camp ($\tilde{N}$), and we assume that, when the infectious population exceeds this capacity, the number of isolated individuals for each class is proportional to the number of clinical symptomatic individuals in the class, i.e. $\tilde{I}_i = \tilde{N} I_i / N_{\mathrm{I}}$. Once this is ascertained, we can encode the different contact rates of these two subpopulations (isolated and not isolated) with other classes in $\lambda_i$. Similar reasoning can be followed regarding the modelization of carers. We do not create a new class since carers are exclusively composed of younger adults with no comorbidities and thus have identical epidemiological parameters to this demographic-class; we only need to modify carers' rate of exposure. Therefore, we can proceed by encoding this intervention in $\lambda_i$ with the parameters $\epsilon_{ij}$ and $\omega_{ij}$ .

Let us start by considering the rate of exposure of younger adults with no comorbidities, hereafter indexed $k$. The general expression of $\lambda_k$ presented in the Main Text is:

$$\lambda_k = \sum_{j=1}^{n} \tau \xi_{kj} c_k \epsilon_{kj} \omega_{kj} \frac{N_j}{N} \left( \frac{\beta_{\mathrm{P}} P_j + \beta_{\mathrm{A}} A_j + \zeta_{\mathrm{I}} \beta_{\mathrm{I}} I_j + \zeta_{\mathrm{H}} \beta_{\mathrm{H}} H_j}{N_j} \right). \tag{24}$$

If we have $N_{\mathrm{care}}$ carers, the expression of $\lambda_k$ can be split in two contributions:

$$\lambda_k = \frac{N_{\mathrm{care}}}{N_k} \lambda_k^{\mathrm{care}} + \left( \frac{N_k - N_{\mathrm{care}}}{N_k} \right) \lambda_k^{\overline{\mathrm{care}}} \tag{25}$$

where the first term in the r.h.s. of the equation is the contribution from carers, and the second term from the remaining population of the class. We can proceed similarly now splitting $\lambda_k^{\mathrm{care}}$ in two, one term to model the interaction with the $\tilde{I}_j$ isolated individuals and another term to account for the interaction with the $I_j - \tilde{I}_j$ not-isolated individuals (if the total number of symptomatic individuals $N_{\mathrm{I}}$ exceeds the capacity of the camp $\tilde{N}$):

$$\lambda_k^{\mathrm{care}} = \tau \sum_{j=1}^{n} \underbrace{\tilde{\xi}_{kj} \tilde{c}_k \overbrace{\tilde{\epsilon}_{kj} \tilde{\omega}_{kj} \frac{N_j}{N} \frac{\tilde{\zeta}_{\mathrm{I}} \tilde{\beta}_{\mathrm{I}} \tilde{I}_j}{N_j}}^{\tilde{P}(k \to j)}}_{\text{isolated}} + \underbrace{\xi_{kj} c_k \epsilon_{kj} \omega_{kj} \frac{N_j}{N} \left( \frac{\beta_{\mathrm{P}} P_j + \beta_{\mathrm{A}} A_j + \zeta_{\mathrm{I}} \beta_{\mathrm{I}} (I_j - \tilde{I}_j) + \zeta_{\mathrm{H}} \beta_{\mathrm{H}} H_j}{N_j} \right)}_{\text{not isolated}}. \tag{26}$$

where we indicate with tildes those parameters in the isolated population and we note that, if these parameters are the same than for the not-isolated population, we retrieve back Eq. 24. The split presented in Eq. 26 is valid under the well-mixed assumption. However, this assumption does not hold for the isolated population, which is confined under very particular conditions. More specifically, we should re-estimate the term highlighted as $\tilde{P}(k \to j)$ which is the probability of interaction of the population of carers with isolated individuals (because, by definition, those fulfilling their roles as carers will interact with isolated individuals), and hence $\tilde{P}(k \to j) = 1$.

Therefore, for the isolated term, we should just estimate the coefficients $\tilde{\xi}_{kj}$ and $\tilde{c}_k$. To estimate $\tilde{c}_k$ we note that, if the class has $N_{\mathrm{care}}$ carers, each isolated individual requires $c_{\mathrm{care}}$ contacts per day with them, and each class $j$ has $\tilde{I}_j$ isolated individuals, the mean number of contacts that each carer has per day with individuals in isolation is $\tilde{c}_k = c_{\mathrm{care}} \sum_j \tilde{I}_j / N_{\mathrm{care}}$. In our simulations, each isolated individual receives one visit per day, i.e. $c_{\mathrm{care}} = 1$. Finally, since we envisage these interactions occuring in what we call buffer zones, namely open spaces in which carers and isolated individuals maintain a distance and wear masks, reducing the transmissibility by 80%, we consider that $\tilde{\xi}_{ij} = 0.2$.

For the not-isolated term, we maintain the well-mixed population assumption explained in the Main Text, with the incorporation of a Heaviside function, $\zeta_I = \Theta(N_{\mathrm{I}} - \tilde{N})$, which activates interaction with clinical symptomatic individuals when their number $N_{\mathrm{I}}$ exceeds the isolation capacity $\tilde{N}$. We should also readjust the probability of interacting with symptomatic non-isolated individuals in class $j$ to be proportional to their fraction of the class' population, $(I_j - \tilde{I}_j)/(N_j - \tilde{I}_j)$. This yields the following expression for the subpopulation of carers:

$$\lambda_k^{\text{care}} = \tau \sum_j \underbrace{\xi_{ij}\beta_{\text{I}}\tilde{c}_k}_{\text{isolated}} + \underbrace{c_k \left(\frac{N_j - \tilde{I}_j}{N}\right)\left(\frac{\beta_{\text{P}}P_j + \beta_{\text{A}}A_j + \beta_{\text{I}}\Theta(N_{\text{I}} - \tilde{N})(I_j - \tilde{I}_j) + \beta_{\text{H}}H_j}{N_j - \tilde{I}_j}\right)}_{\text{not isolated}}. \tag{27}$$

Note that the mean number of contacts per day that carers have with the rest of the population, $c_k$, is not reduced since we do not make additional assumptions about carers' behavioural changes outside of their role as carers.

Younger adults without comorbidities who do not serve as carers will not interact with isolated individuals (i.e. $\tilde{P}(k \to j) = 0$), and hence their rate of exposure becomes:

$$\lambda_k^{\overline{\text{care}}} = \underbrace{c_k \left(\frac{N_j - \tilde{I}_j}{N}\right)\left(\frac{\beta_{\text{P}}P_j + \beta_{\text{A}}A_j + \beta_{\text{I}}\Theta(N_{\text{I}} - \tilde{N})(I_j - \tilde{I}_j) + \beta_{\text{H}}H_j}{N_j - \tilde{I}_j}\right)}_{\text{not isolated}}. \tag{28}$$

Inserting Eqs. 27 and 28 into Eq. 25 yields:

$$\lambda_k = \tau \sum_j \underbrace{\xi_{ij}\beta_{\text{I}}c_{\text{care}}\frac{\tilde{I}_j}{N_k}}_{\text{isolated}} + \underbrace{c_k \left(\frac{\beta_{\text{P}}P_j + \beta_{\text{A}}A_j + \beta_{\text{I}}\Theta(N_{\text{I}} - \tilde{N})(I_j - \tilde{I}_j) + \beta_{\text{H}}H_j}{N}\right)}_{\text{not isolated}}. \tag{29}$$

We can express this equation following the parameterization presented in Eq. 24 making $\tilde{\xi}_{ij} = 0.2$, $\tilde{\epsilon}_{ij} = (c_{\text{care}}/c_i)(\tilde{I}_j/N_i)$, $\tilde{\omega}_{ij} = N/N_j$ and $\tilde{\zeta}_{\text{I}}\tilde{\beta}_{\text{I}} = 1$ for $j \in$ isolated individuals in class $j$, while $\xi = 1$,$\epsilon_{ij} = 1$, $\omega_{ij} = 1$ and $\zeta_{\text{I}} = \Theta(N_{\text{I}} - \tilde{N})$ for $j \in$ individuals in class $j$ who are not isolated. The rate of exposure of the remaining classes (not younger adults without comorbidities, $i \neq k$) corresponds to the second term in the r.h.s of Eq. 29:

$$\lambda_i = \tau \sum_j \underbrace{c_i \frac{\beta_{\text{P}}P_j + \beta_{\text{A}}A_j + \beta_{\text{I}}\Theta(N_{\text{I}} - \tilde{N})(I_j - \tilde{I}_j) + \beta_{\text{H}}H_j}{N}}_{\text{not isolated}}. \tag{30}$$

We should note that Eq. 29 does not depend on the number of carers, $N_{\text{care}}$, since we assume that the rate of exposure of carers is evenly distributed among all individuals in the class of healthy younger adults. If this assumption were not made, a specific class of carers could be created and the variable $N_{\text{care}}$ maintained explicit.

#### 3 Supplementary figures

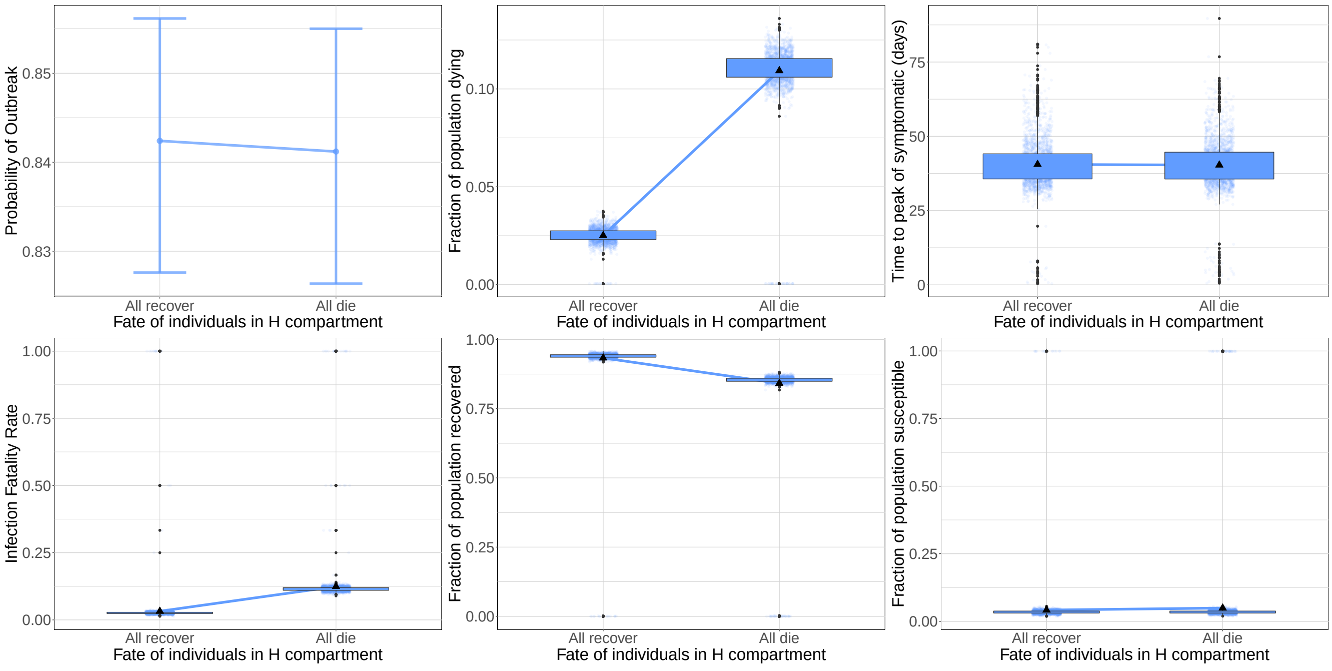


Figure 1: **Outcomes when all severe (hospitalized) cases recover ($\sigma = 0$) vs when all severe (hospitalized) cases die ($\sigma = 1$).** Probability of an outbreak (top left), fraction of the population dying (top middle), time until peak symptomatic cases (top right), IFR (bottom left), and fraction of the population that recovers (bottom middle). Since we define outbreaks as simulations in which at least one person dies and probability of a case dying is higher when $\sigma = 1$, the probability of observing an outbreak is also necessarily higher when $\sigma = 1$.

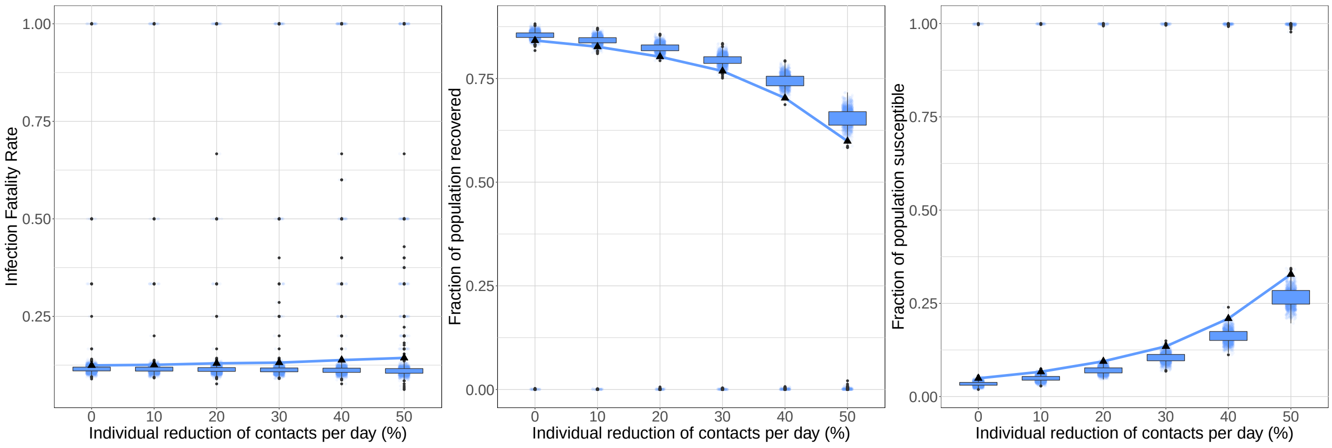


Figure 2: **Self-distancing.** IFR (left), and fraction of the population that recovers (right) as a function of the proportion of contacts reduced per individual per day.

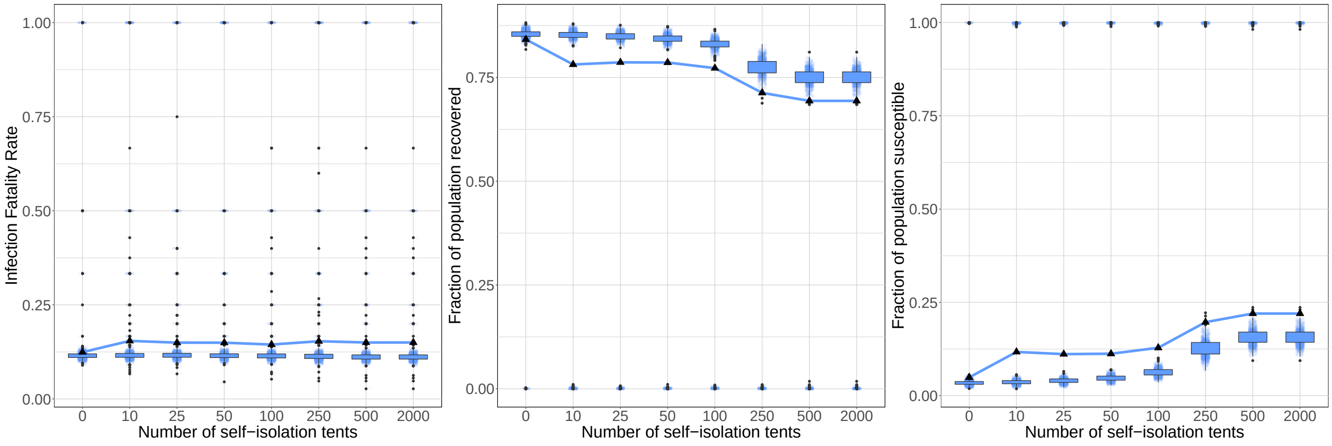


Figure 3: **Self-isolation.** IFR (left), and fraction of the population that recovers (right) as a function of the number of isolation tents available in the camp.

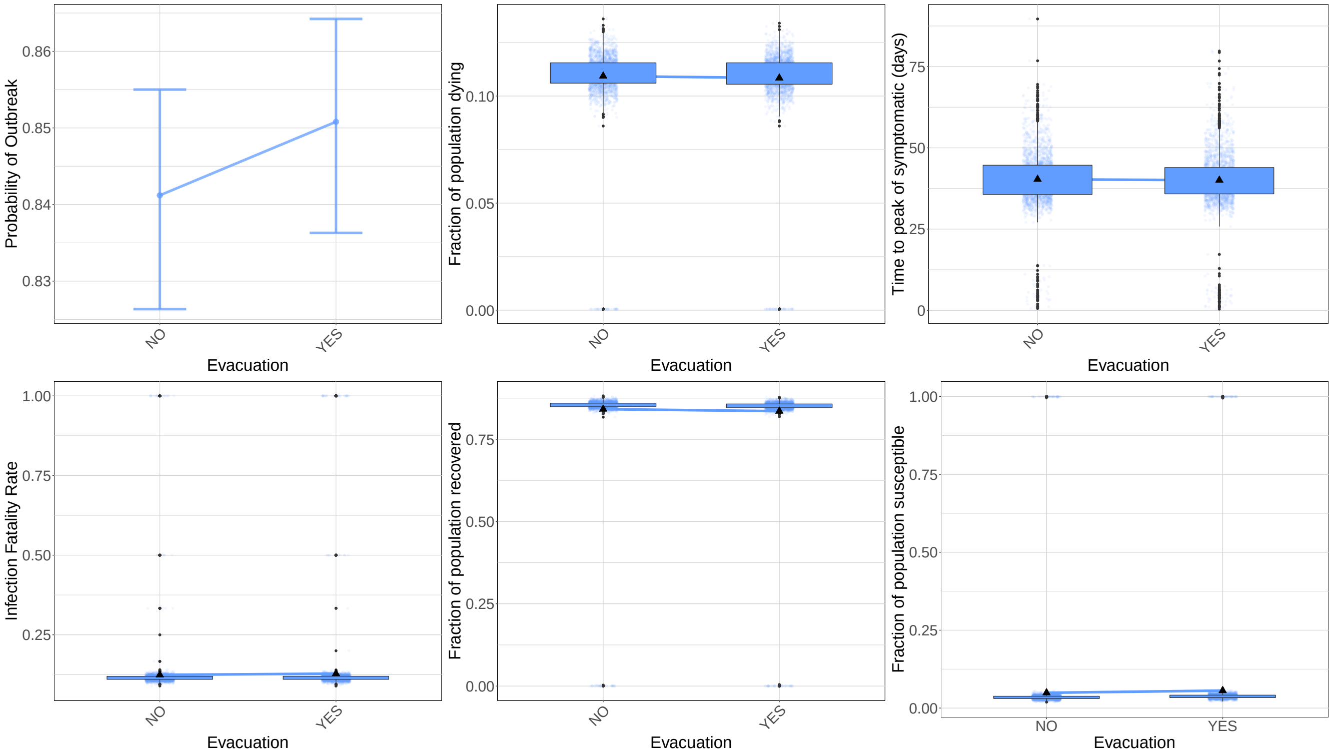


Figure 4: **Evacuation.** Probability of an outbreak (top left), fraction of the population dying (top middle), time until peak symptomatic cases (top right), IFR (bottom left), and fraction of the population that recovers (bottom middle), as a function of whether individuals requiring hospitalization are evacuated to isolation centers.

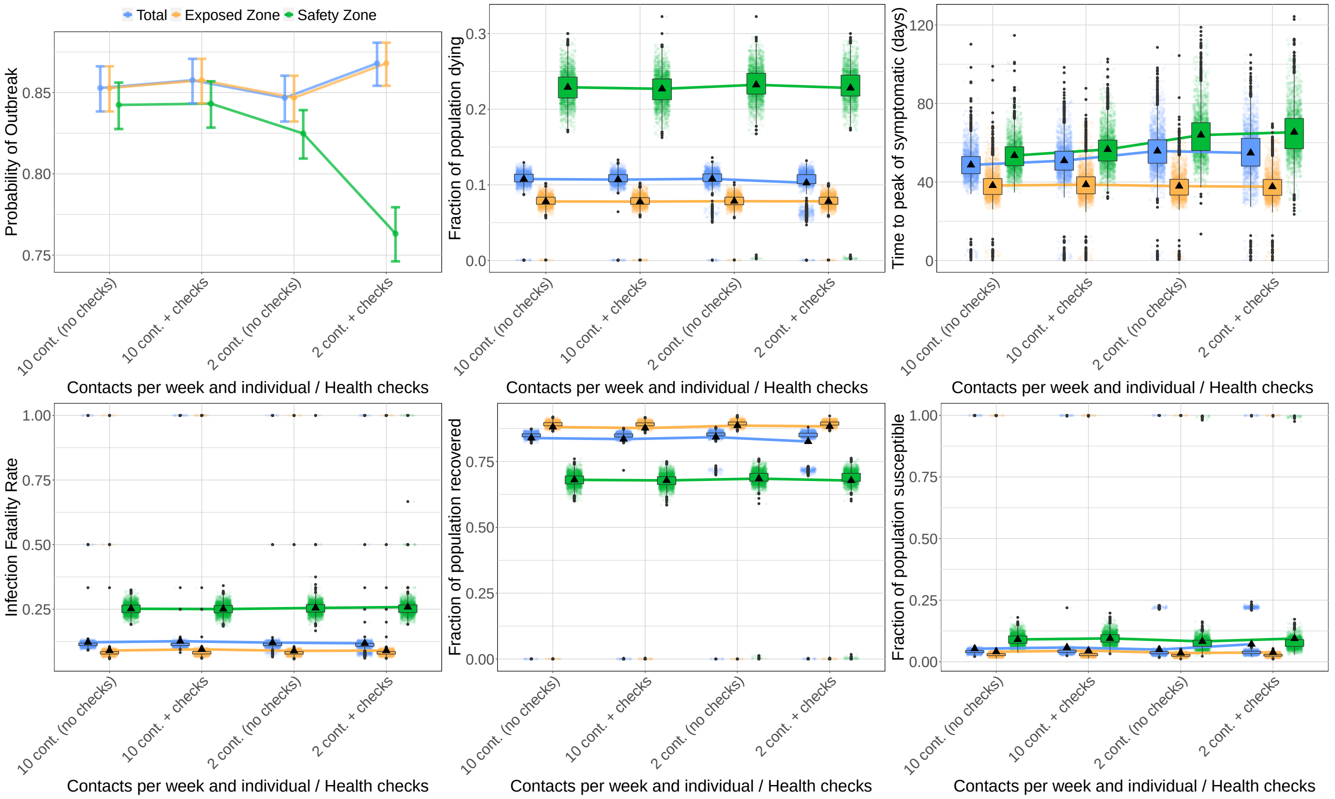


Figure 5: **Health-checks in the buffer zone.** Probability of an outbreak (top left), fraction of the population dying (top middle), time until peak symptomatic cases (top right), IFR (bottom left), and fraction of the population that recovers (bottom middle), as a function of whether health-checks are implemented in the buffer zone between the safety and exposed zones. Scenarios with 10 or 2 contacts in the buffer zone per person in the safety zone per week are plotted. All figures consider the scenario in which 20% of the camp's population is allocated to the safety zone. Note that the mean of an outcome for the whole population is not the weighted mean of the exposed and safety zones, since outcomes are computed considering simulations in which at least one death was observed in the population class inhabiting the zone, i.e. the number of simulations considered to compute each mean may be different. This explains for example why there is a reduction in the mean time until symptomatic cases peak when moving from 2 contacts per week without health checks to considering health checks for the whole population, despite there being an increase in the safety zone.

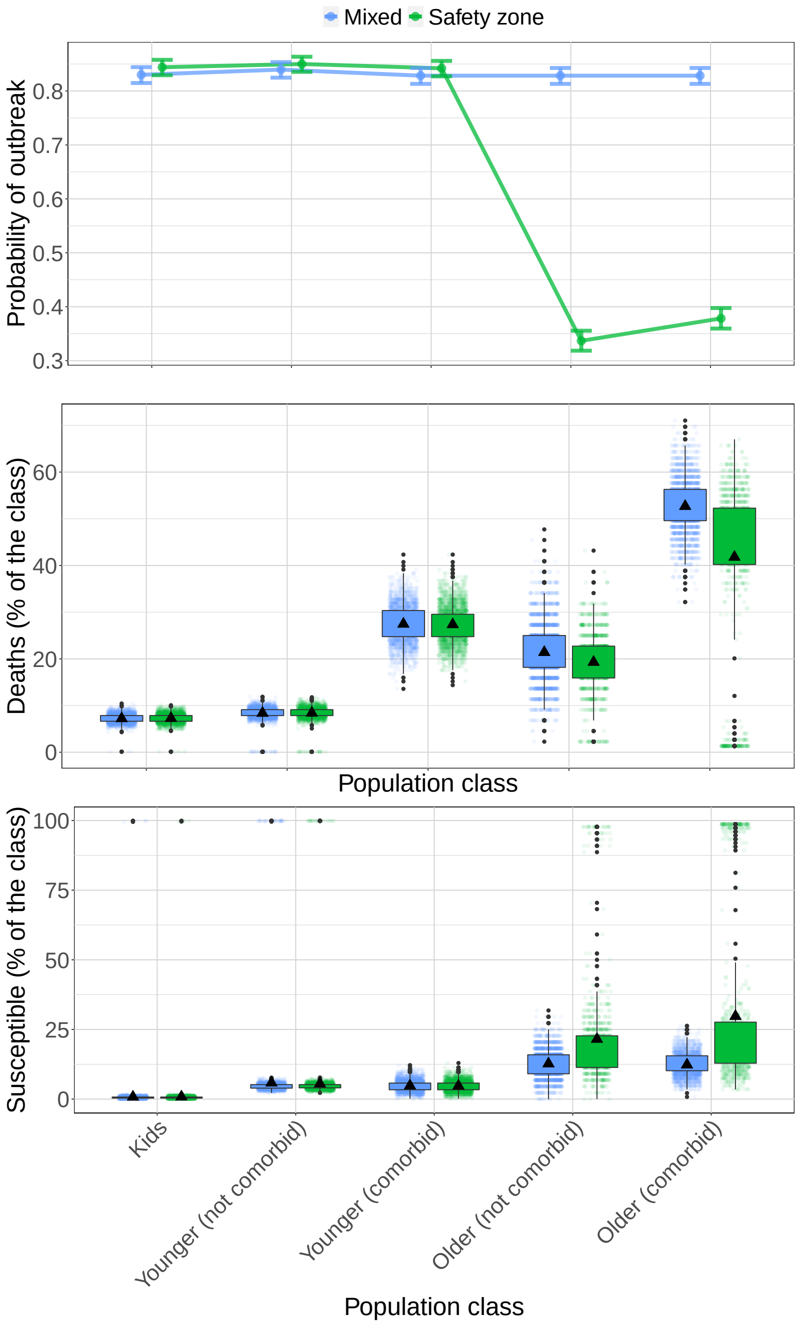


Figure 6: **Effects of the safety zone on outcomes by population class.** Probability of an outbreak (top), and proportion that dies in each population class (bottom) when no interventions are implemented (Mixed), compared to protection of older adults in the safety zone with 2 contacts in the buffer zone per week (Safety zone). The fraction of deaths in the safety zone for the older population is significantly lower.

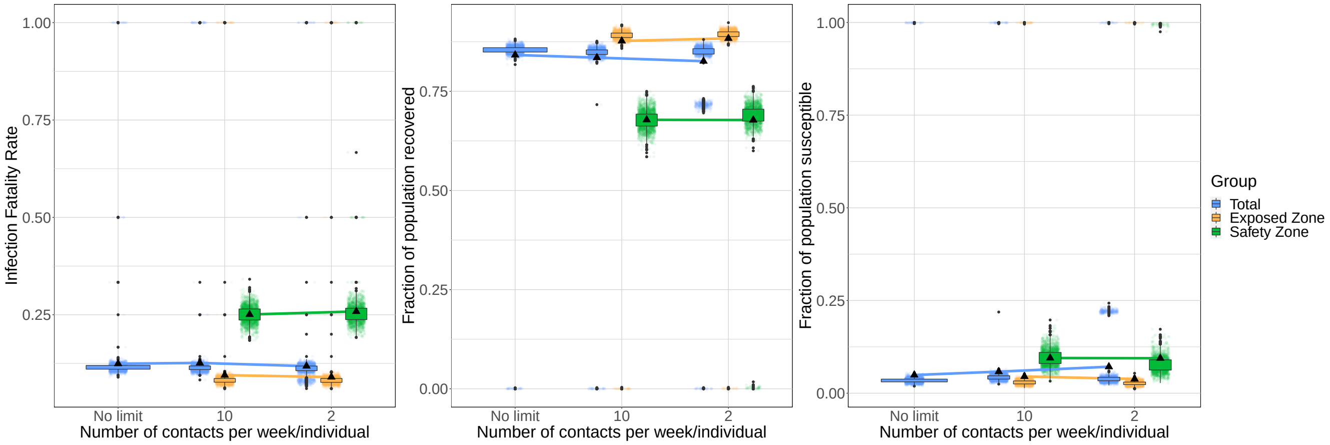


Figure 7: **Number of contacts in the buffer zone.** IFR (left), and fraction of the population that recovers (right) as a function of the number of contacts that each individual in the safety zone has in the buffer zone per week. All figures consider the scenario in which 20% of the camp's population is allocated to the safety zone.

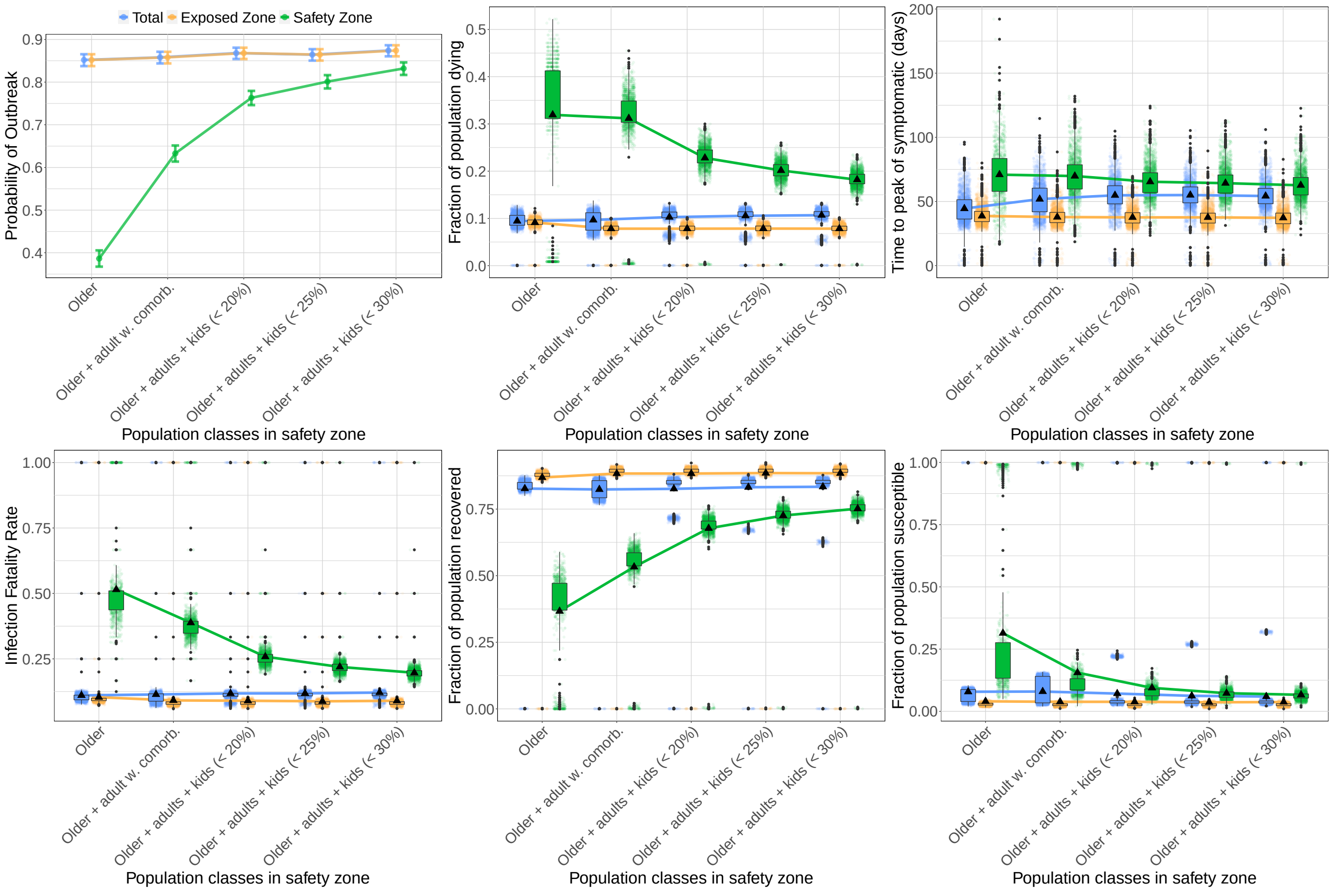


Figure 8: **Population moving to the safety zone.** Probability of an outbreak (top left), fraction of the population dying (top middle), time until peak symptomatic cases (top right), IFR (bottom left), and fraction of the population that recovers (bottom middle) as a function of the safety zone allocation scenario (see Table 2). All figures consider the scenario with 2 contacts in the buffer per person in the safety zone per week.

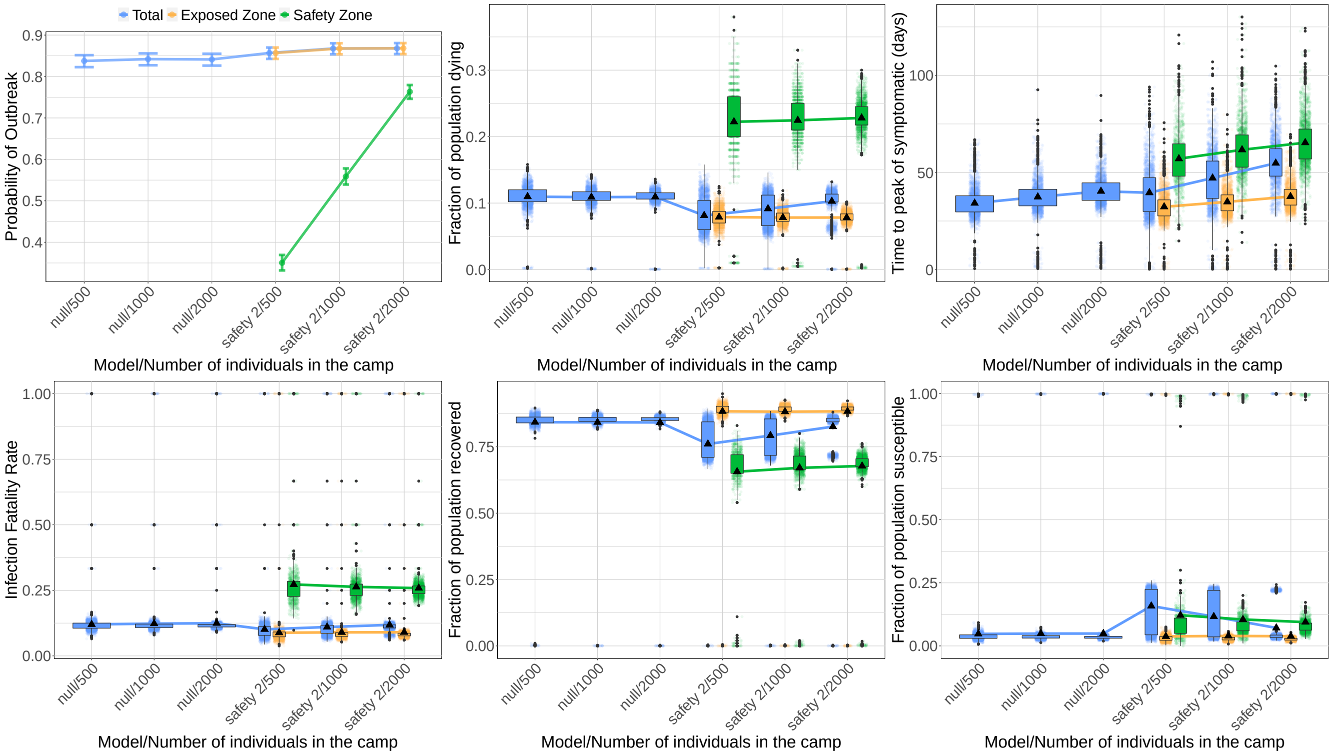


Figure 9: **Efficacy of the safety zone for different population sizes.** Probability of an outbreak (top left), fraction of the population dying (top middle), time until peak symptomatic cases (top right), IFR (bottom left), and fraction of the population that recovers (bottom middle) as a function of the total population size. The figures consider scenarios with no interventions (null), and with a safety zone comprising 20% of the camp's population with 2 contacts in the buffer zone per person in the safety zone per week (safety 2).

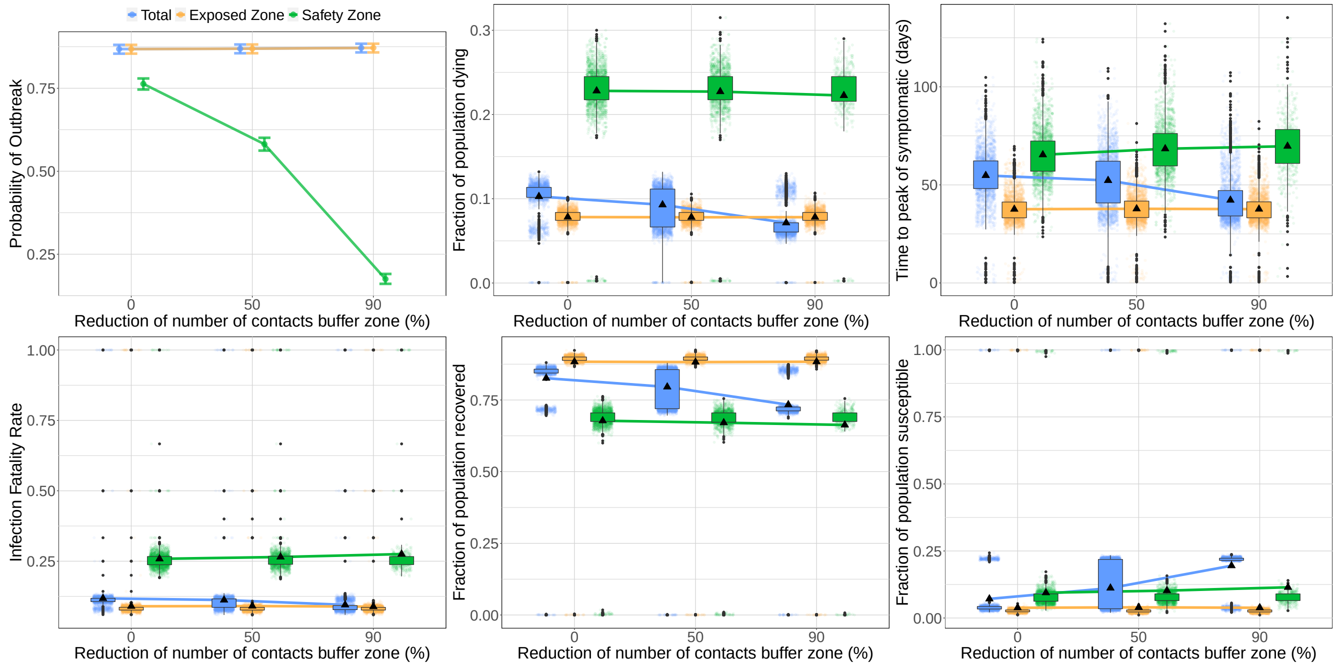


Figure 10: **Lockdown of the safety zone.** Probability of an outbreak (top left), fraction of the population dying (top middle), time until peak symptomatic cases (top right), IFR (bottom left), and fraction of the population that recovers (bottom middle) as a function of the reduction in the number of contacts permitted in the buffer zone from a baseline of 2 per person in the safety zone per week. All figures consider the scenario in which 20% of the camp's population is allocated to the safety zone.

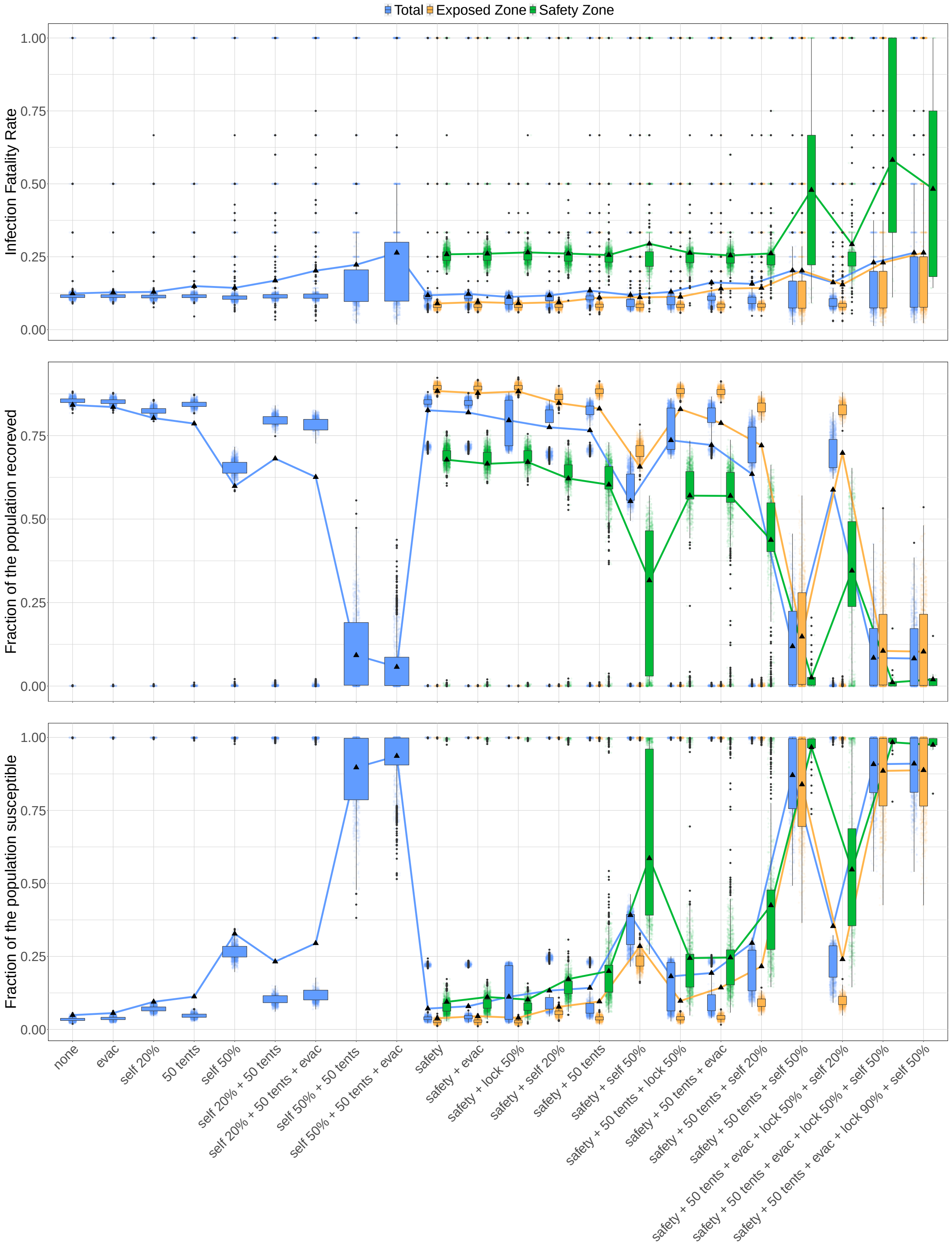


Figure 11: **Combined interventions.** IFR (top), and fraction of the population that recovers (bottom) for different combinations of interventions. Evac = evacuation of severely symptomatic, self = self-distancing, tents = number of available self-isolation tents, safety = safety zone, lock = lockdown of the buffer zone. For combinations of interventions including a safety zone, we distinguish between the population living in the green zone, in the orange zone and the whole population. The increase in the IFR for the green zone is explained by the discretization of the possible values that the IFR can take when the number of cases is very low (see Supplementary Table 3).

| Intervention | <20 cases | Total | % of total |
|---|---|---|---|
| safety | 36 | 1908 | 1.9 |
| safety + evac | 48 | 1894 | 2.5 |
| safety + lock 50% | 40 | 1454 | 2.8 |
| safety + self 20% | 56 | 1582 | 3.5 |
| safety + 50 tents | 37 | 1330 | 2.8 |
| safety + self 50% | 187 | 719 | 26 |
| safety + 50 tents + lock 50% | 58 | 1021 | 5.7 |
| safety + 50 tents + evac | 44 | 1245 | 3.5 |
| safety + 50 tents + self 20% | 78 | 907 | 8.6 |
| safety + 50 tents + self 50% | 57 | 71 | 80 |
| safety + 50 tents + evac + lock 50% + self 20% | 108 | 594 | 18 |
| safety + 50 tents + evac + lock 50% + self 50% | 42 | 44 | 95 |
| safety + 50 tents + evac + lock 90% + self 50% | 14 | 15 | 93 |

Table 3: **Efficacy of the safety zone in combination with other interventions.** <20 cases = number of outbreaks in the green zone with fewer than 20 cases recorded. Total = total number of simulations where an outbreak in the green zone occurs (at least one death). % of total = percent of outbreaks where fewer than 20 cases are recorded. N = 2500 simulations for each combination of interventions. For the most effective combinations, the majority of simulations where an outbreak occurs in the green zone see fewer than 20 cases. In these simulations, the discretization of the possible values that the IFR can take explains its apparently anomalous increase in Fig. 11.

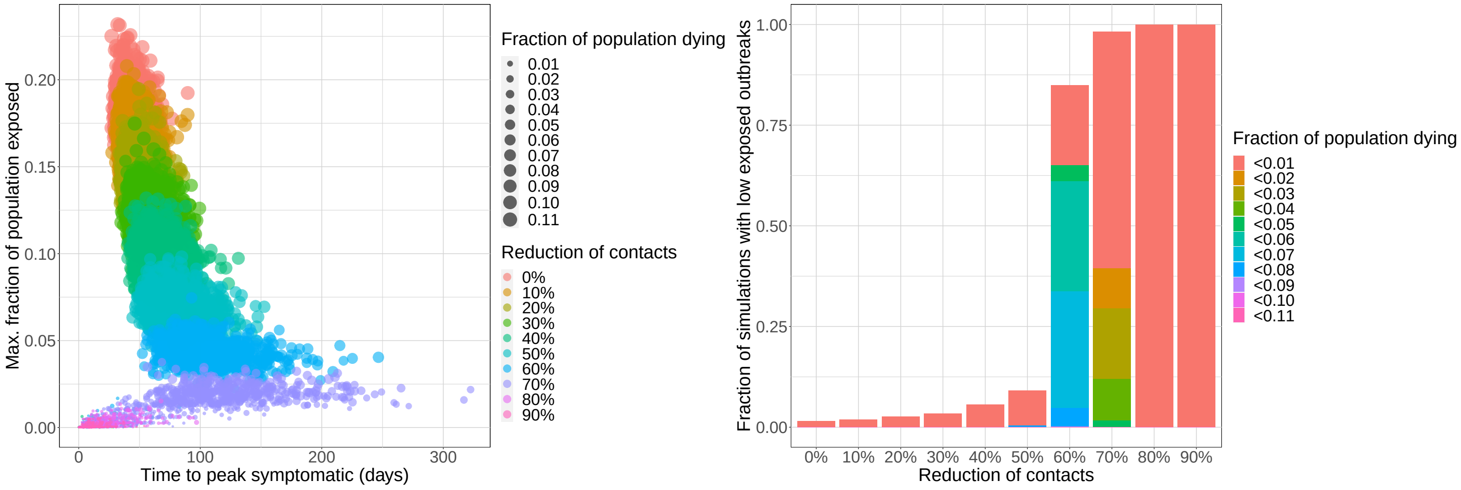


Figure 12: **Critical number of exposed individuals.** (Left) Reducing the number of contacts reduces the maximum number of individuals simultaneously exposed while increasing the time until symptomatic cases peak. When the reduction goes beyond 60%, there are abrupt drops in the time until cases peak and the fraction of the population dying, suggesting there is a critical number of individuals who must be exposed, under which outbreaks die out before spreading widely throughout the population. Above this threshold, the virus becomes established in the population over a longer period of time, increasing mortality. (Right) Fraction of the simulations which do not achieve the critical number of exposed individuals (that we set to 2% of the population). The abrupt transition between 50 and 70% reductions in contacts is apparent.

### 4 Appendix: List of Experiments

| Simul. | Experim. | Structure | Npop | Evac. | N.Tents | Contacts | Tcheck | lock | self | H-fate |
|---|---|---|---|---|---|---|---|---|---|---|
| 1 | A. Shielding and limits | Null Mixed | 2000 | No | 0 | MF | No | No | No | D |
| 2 | | | | | | | | | | R |
| 3 | | Shield (20%) | | | | 2/7 | Yes | | | D |
| 4 | | | | | | | | | | R |
| 5 | | | | | | 10/7 | | | | D |
| 6 | | | | | | | | | | R |
| 7 | B. Role individual tents | Null Mixed | 2000 | No | 10 | MF | No | No | No | D |
| 8 | | | | | 20 | | | | | |
| 9 | | | | | 50 | | | | | |
| 10 | | | | | 100 | | | | | |
| 11 | | | | | 250 | | | | | |
| 12 | | | | | 500 | | | | | |
| 13 | | | | | ∞ | | | | | |
| 14 | | Shield (20%) | | | 10 | 2/7 | Yes | | | |
| 15 | | | | | 20 | | | | | |
| 16 | | | | | 50 | | | | | |
| 17 | | | | | 100 | | | | | |
| 18 | | | | | 250 | | | | | |
| 19 | | | | | 500 | | | | | |
| 20 | | | | | ∞ | | | | | |
| 21 | C. Population shielded | Shield (age3) | 2000 | No | 0 | 2/7 | Yes | No | No | D |
| 22 | | Shield (age2) | | | | | | | | |
| 23 | | Shield (20%) | | | | | | | | |
| 24 | | Shield (25%) | | | | | | | | |
| 25 | | Shield (30%) | | | | | | | | |
| 26 | D. Population size | Null Mixed | 500 | No | 0 | MF | No | No | No | D |
| 27 | | | 1000 | | | | | | | |
| 28 | | Shield (20%) | 500 | | | 2/7 | Yes | | | |
| 29 | | | 1000 | | | | | | | |
| 30 | E. Safety zone testing | Shield (20%) | 2000 | No | 0 | 2/7 | No | No | No | D |
| 31 | | | | | | | Yes | | | |
| 32 | | | | | | 10/7 | No | | | |
| 33 | | | | | | | Yes | | | |

Table 4: **List of simulations performed.** Npop = Population size. Evac. = Is people requiring hospitalization evacuated? N. tents = Number self-isolation tents per camp. Contacts = Number of contacts per day between populations shielded. Tcheck = Are temperature checks performed? Lock = Is lockdown applied after first symptomatic case is identified? self = Fraction of contacts remaining after self-distancing is implemented. H-fate = Final compartment for hospitalized people. MF = Mean field. Shield = Population shielded. age3 = elderly population. age2 = adults with comorbidities and spouses. (20-30%) = kids from adults shielded up to x% of total population.

| Simul. | Experim. | Structure | Npop | Evac. | N. tents | Contacts | Tcheck | lock | self | H-fate |
|---|---|---|---|---|---|---|---|---|---|---|
| 34 | F. Lockdown | Shield (20%) | 2000 | No | 0 | 2/7 | Yes | 0.5 | No | D |
| 35 | | | | | | | | 0.9 | | |
| 36 | | | | | | | | 0.99 | | |
| 37 | G. Self-distancing | Null Mixed | 2000 | No | 0 | MF | No | No | 0.1 | D |
| 38 | | | | | | | | | 0.2 | |
| 39 | | | | | | | | | 0.3 | |
| 40 | | | | | | | | | 0.4 | |
| 41 | | | | | | | | | 0.5 | |
| 42 | | Shield (20%) | | | | 2/7 | Yes | | 0.1 | |
| 43 | | | | | | | | | 0.2 | |
| 44 | | | | | | | | | 0.3 | |
| 45 | | | | | | | | | 0.4 | |
| 46 | | | | | | | | | 0.5 | |
| 47 | H. Evacuation | Null Mixed | 2000 | Yes | 0 | MF | No | No | No | D |
| 48 | | Shield (20%) | | | | 2/7 | Yes | | | |
| 49 | I. Combined interventions | Null Mixed | 2000 | No | 50 | MF | No | No | 0.2 | D |
| 50 | | | | | | | | | 0.5 | |
| 51 | | Shield (20%) | | | | 2/7 | Yes | No | 0.2 | |
| 52 | | | | | | | | 0.5 | No | |
| 53 | | | | | | | | 0.5 | 0.2 | |
| 54 | | | | | | | | 0.9 | 0.2 | |
| 55 | | | | | | | | 0.5 | 0.5 | |
| 56 | | | | | | | | 0.9 | 0.5 | |
| 57 | | Null Mixed | 2000 | Yes | 50 | MF | No | No | 0.2 | D |
| 58 | | | | | | | | | 0.5 | |
| 59 | | Shield (20%) | | | | 2/7 | Yes | No | 0.2 | |
| 60 | | | | | | | | 0.5 | No | |
| 61 | | | | | | | | 0.5 | 0.2 | |
| 62 | | | | | | | | 0.9 | 0.2 | |
| 63 | | | | | | | | 0.5 | 0.5 | |
| 64 | | | | | | | | 0.9 | 0.5 | |
| 65 | | | 500 | | | | | No | 0.2 | |
| 66 | | | | | | | | 0.5 | No | |
| 67 | | | | | | | | 0.5 | 0.2 | |

Table 5: **List of simulations performed (II).** Npop = Population size. Evac. = Is people requiring hospitalization evacuated? N. tents = Number self-isolation tents per camp. Contacts = Number of contacts per day between populations shielded. Tcheck = Are temperature checks performed? Lock = Is lockdown applied after first symptomatic case is identified? self = Fraction of contacts remaining after self-distancing is implemented. H-fate = Final compartment for hospitalized people. MF = Mean field. Shield = Population shielded. age3 = elderly population. age2 = adults with comorbidities and spouses. (20-30%) = kids from adults shielded up to x% of total population.

[2] Peter Ashcroft, Jana S. Huisman, Sonja Lehtinen, Judith A. Bouman, Christian L. Althaus, Roland R. Regoes, and Sebastian Bonhoeffer. COVID-19 infectivity profile correction. *arXiv:2007.06602 [q-bio, stat]*, July 2020. arXiv: 2007.06602.

[9] Jeroen J.A. van Kampen, David A.M.C. van de Vijver, Pieter L.A. Fraaij, Bart L. Haagmans, Mart M. Lamers, Nisreen Okba, Johannes P.C. van den Akker, Henrik Endeman, Diederik A.M.P.J. Gommers, Jan J. Cornelissen, Rogier A.S. Hoek, Menno M. van der Eerden, Dennis A. Hesselink, Herold J. Metselaar, Annelies Verbon, Jurriaan E.M. de Steenwinkel, Georgina I. Aron, Eric C.M. van Gorp, Sander van Boheemen, Jolanda C. Voermans, Charles A.B. Boucher, Richard Molenkamp, Marion P.G. Koopmans, Corine Geurtsvankessel, and Annemiek A. van der Eijk. Shedding of infectious virus in hospitalized patients with coronavirus disease-2019 (COVID-19): duration and key determinants. *medRxiv*, page 2020.06.08.20125310, January 2020.

[10] Oyungerel Byambasuren, Magnolia Cardona, Katy Bell, Justin Clark, Mary-Louise McLaws, and Paul Glasziou. Estimating the extent of asymptomatic COVID-19 and its potential for community transmission: Systematic review and meta-analysis. *Official Journal of the Association of Medical Microbiology and Infectious Disease Canada*, 5(4):223–234, December 2020.

[11] O. Diekmann, Hans Heesterbeek, and Tom Britton. *Mathematical tools for understanding infectious diseases dynamics*. Princeton series in theoretical and computational biology. Princeton University Press, Princeton, 2013.

[12] Sydney Philipps, Dan Rossi, Rachel Von Arb, and Alex Capaldi. Mathematical models of infectious diseases: Two-strain infections in metapopulations, July 2011.

[13] Marino Gatto, Enrico Bertuzzo, Lorenzo Mari, Stefano Miccoli, Luca Carraro, Renato Casagrandi, and Andrea Rinaldo. Spread and dynamics of the COVID-19 epidemic in Italy: Effects of emergency containment measures. *Proceedings of the National Academy of Sciences*, 117(19):10484–10491, May 2020.

[14] J.M Heffernan, R.J Smith, and L.M Wahl. Perspectives on the basic reproductive ratio. *Journal of the Royal Society Interface*, 2(4):281–293, sep 2005.

[15] Yuanyuan Dong, Xi Mo, Yabin Hu, Xin Qi, Fang Jiang, Zhongyi Jiang, and Shilu Tong. Epidemiological characteristics of 2143 pediatric patients with 2019 coronavirus disease in china. *Pediatrics*, 2020.

[16] Nancy Chow, Katherine Fleming-Dutra, Ryan Gierke, Aron Hall, Michelle Hughes, Tamara Pilishvili, Matthew Ritchey, et al. Preliminary estimates of the prevalence of selected underlying health conditions among patients with coronavirus disease 2019 in USA, february 12–march 28, 2020. *Morbidity and Mortality Weekly Report*, 69(13):382, 2020.
